# Age-Specific All-Cause Mortality Disparities by Race and Ethnicity During the COVID-19 Pandemic

**DOI:** 10.1101/2024.03.01.24303592

**Authors:** Jeremy Samuel Faust, Benjamin Renton, Tasce Bongiovanni, Alexander Junxiang Chen, Karen Dorsey Sheares, Chengan Du, Utibe R. Essien, Elena Fuentes-Afflick, Trent Haywood, Rohan Khera, Terris King, Shu-Xia Li, Zhenqiu Lin, Yuan Lu, Andrew D.A. Marshall, Chima D. Ndumele, Ijeoma Opara, Tina Loarte-Rodriguez, Mitsuaki Sawano, Kekoa Taparra, Herman A. Taylor, Karol E. Watson, Clyde W. Yancy, Harlan M. Krumholz

## Abstract

**BACKGROUND:** The end of the public health emergency provides an opportunity to fully describe disparities during the Covid-19 pandemic.

**METHODS:** In this retrospective cohort analysis of US deaths during the Covid-19 public health emergency (March 2020-April 2023), all-cause excess mortality and years of potential life lost (YPLL) were calculated by race or ethnicity overall and by age groups (ages <25 years, 25-64 years, ≥65 years). Temporal correlations with Covid-19-specific mortality were measured.

**RESULTS:** >1.38 million all-cause excess deaths and ~23 million corresponding YPLL occurred during the pandemic. Had the rate of excess mortality observed among the White population been observed among the total population, >252,300 (18%) fewer excess deaths, and >5,192,000 fewer (22%) YPLL would have occurred. The highest excess mortality rates were among the American Indian/Alaska Native (AI/AN, 822 per 100,000; ~405,700 YPLL) and the Black (549 per 100,000; ~4,289,200 YPLL) populations. The highest relative increase in mortality was observed in the AI/AN population (1.34; 95% CI 1.31-1.37), followed by Hispanic (1.31; 95% CI 1.27-1.34), Native Hawaiian or Other Pacific Islander (1.24; 95% CI 1.21-1.27), Asian (1.20; 95% CI 1.18-1.20), Black (1.20; 95% CI 1.18-1.22) and White (1.12; 95% CI 1.09-1.15) populations. Greater disparities occurred among children and adults <65 years.

**CONCLUSIONS:** Excess mortality occurred in all groups during the Covid-19 pandemic, with disparities by race and ethnicity, especially in younger and middle-aged populations. >252,000 and 5.2 million fewer YPLL would have been observed had increases in mortality among the total population been similar to the White population.

## Introduction

The end of the Covid-19 public health emergency in the US provides an opportunity to evaluate mortality disparities. A focus on all-cause mortality allows a comprehensive perspective of the overall impact of the pandemic, capturing direct Covid-19 deaths (including miscategorized Covid-19-related deaths) and indirect consequences of the pandemic, including consequences for populations that are frequently excluded from public-facing reports and dashboards.^1–8^

We assessed mortality disparities by race and ethnicity during the Covid-19 pandemic, overall and stratified by age, during the US public health emergency. Using CDC data, we modeled and measured cumulative all-cause excess mortality, analyzed the age distribution of excess deaths, and estimated the years of potential lost life (YPLL) for each race and ethnicity, for age groups, and determined how many of these deaths and years of lost life could have been averted, had subgroup ratios replicated those observed in the White population. To determine whether existing disparities were replicated or augmented during the pandemic, pre-pandemic and pandemic all-cause mortality risks by race/ethnicity and age were measured, with attention to variation during Covid-19 waves and differences between the pre-vaccine and vaccine eras.

## Methods

To measure pandemic-associated mortality by race/ethnicity, we adapted established methods for computing age-adjusted rates, estimating excess mortality and years of potential life lost.^1,5–10^ Using the CDC Single Race designations, we divided the US population into seven race/ethnicity groups: Non-Hispanic American Indian/Alaska Native (AI/AN), Non-Hispanic Asian (Asian), Non-Hispanic-Black or African American (Black), Hispanic of all races (Hispanic), Non-Hispanic Native Hawaiian or Other Pacific Islander (NHPI), and Non-Hispanic White (White). Persons who reported “more than one race” were included for completeness but not analyzed as a group.^11^ Hispanic ethnicity was not stated for 0.0027% of deaths and these deaths were excluded (race categorization was available for all deaths). We divided each race/ethnicity into three age groups: <25, 25-64, and ≥65 years. The 38-month study period was March 1, 2020-April 30, 2023.

## Data

We used deidentified, publicly available death and US Census population data from the CDC’s Wide-ranging Online Data for Epidemiologic Research (WONDER), which encompasses all deaths among US residents.^12^

## Analyses

### Excess Mortality

Excess mortality was defined as the number of raw, observed, deaths minus modeled expected deaths.^9,10^ To estimate expected deaths, we used seasonal, autoregressive integrated moving averages (ARIMA) for all-cause mortality by component demographic (race/ethnicity and age grouping). For each demographic, ARIMA models were trained using pre-pandemic population data (2014-2020) to estimate yearly populations for 2021-2023 (estimated yearly changes were divided evenly over 12 months). Seasonal ARIMA models were trained on monthly death counts (62 pre-pandemic months, January 2015-February 2020) for each age group, using the estimated monthly populations as covariates to overcome stationarity. Composite all-ages estimates were summed from the component age-group models and populations were corrected for cumulative excess mortality (see Supplement, p.2-3).

### Years of Potential Life Lost

To estimate total and per capita years of potential life lost (YPLL), we modeled excess deaths for each race/ethnicity by 10-year age group (except <25 years and >85 years) and applied CDC life table figures to calculate the expectation of life for single year ages for each race/ethnicity (see Supplement, p.3).^13^

### Potential Excess Deaths and YPLL Averted

Potential excess deaths and YPLL averted were calculated by applying the observed-to-expected ratios observed in the same-age White population to each race/ethnicity group and summed for overall tallies (See Supplement, p.3).

### Disparity Rate Ratios

We calculated disparity rate ratios (DRR) during the pandemic by dividing the share of the excess mortality and YPLL of each race/ethnicity, within each age group, by the population share within that age group. 95% CIs were determined using bandwidths from the excess mortality model (See Supplement, p.4).

### Cause-Specific Excess Mortality

An analysis of cause-specific excess mortality was conducted to characterize disparities by specific medical (i.e., “natural”) causes and external manner of death (e.g., accidents, homicide). We applied the methods described above, with two modifications based on data availability (see Supplement, p.4). Pearson coefficients correlating all-cause excess mortality and select medical causes of death (including Covid-19; ICD U00-U99: Codes for special purposes, of which Covid-19 comprised >99.992% of deaths) and for external manner of death, were determined to assess temporal associations. Correlations ≥0.6 were considered strong; 0.4-0.59 were considered moderate.

### Changes in Mortality Disparities

To determine whether pre-pandemic mortality disparities changed over the course of the pandemic, we measured pre-pandemic (March 2015-February 2020) and pandemic relative risks (RR) for all-cause mortality incidence rates for every group and used the contemporaneous same-age White population as comparators. Yearly and monthly RRs were determined for pre-pandemic (e.g., March 2015-February 2016) and pandemic years (e.g., March 2020-February 2021). 95% confidence intervals for all RRs were determined via the geometric means of monthly RRs within specific periods (yearly, pre-pandemic, and pandemic).

Analyses were performed in R 4.0.3 and Microsoft Excel 16.74. Exhibits were created in Flourish (Canva UK Operations, London, UK). This study of de-identified publicly available data complied with the GATHER checklist and did not require institutional review board approval.^14^

## Results

### Excess Deaths and Years of Potential Lost Life

There were >1.38 million all-cause excess deaths and ~23 million corresponding years of potential life lost (YPLL) during the pandemic. Had the rate of excess mortality observed among the White population been observed among the total population, >252,300 (18%) fewer excess deaths, corresponding to >5,192,000 fewer (22%) YPLL would have occurred, including >133,800 fewer excess deaths and >2,899,600 YPLL in the Hispanic population alone. (Table 1).

**Table 1.**
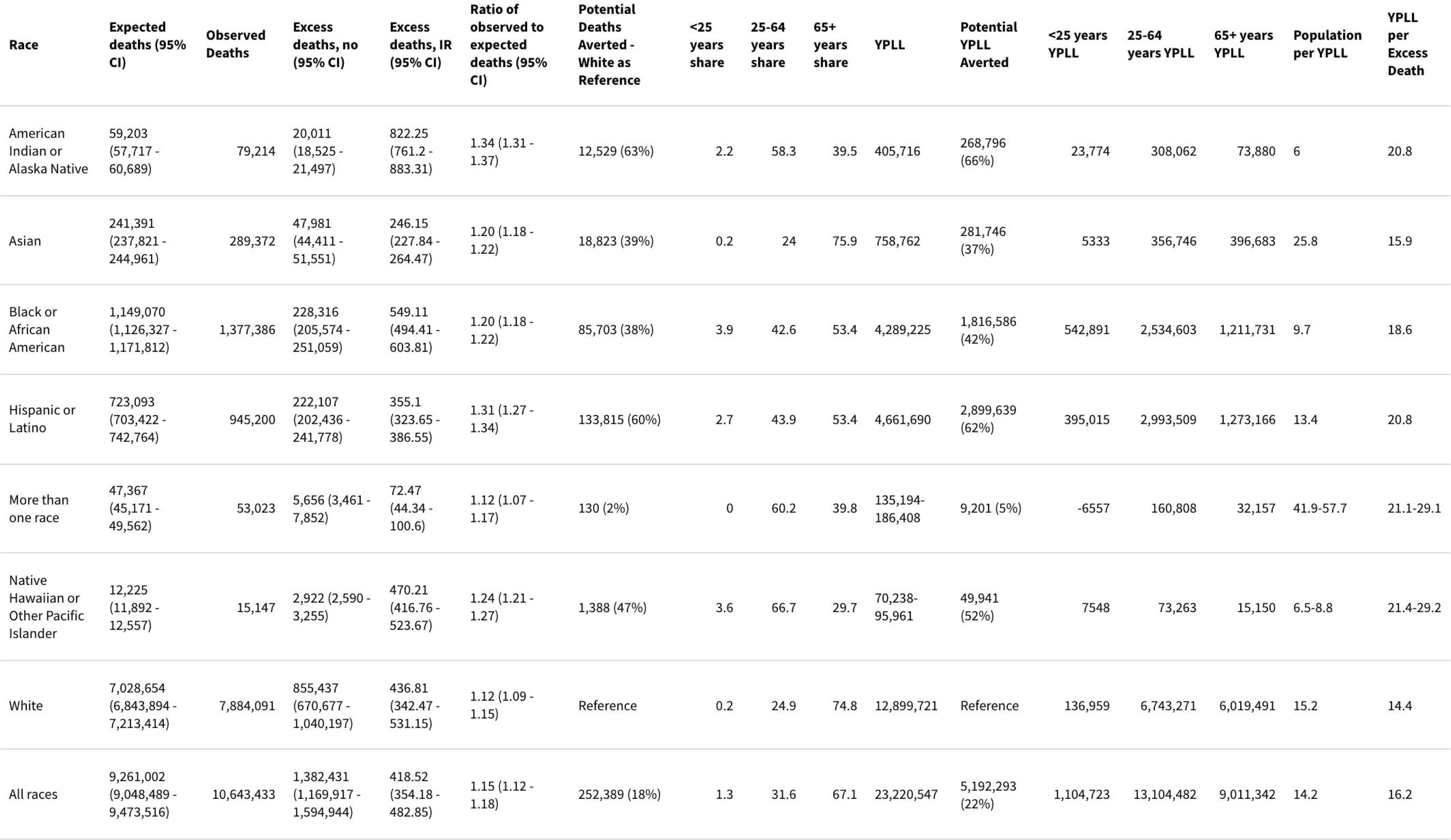
All-cause mortality, excess deaths, potential excess deaths averted, years of potential life lost (YPLL), and potential YPLL averted by race/ethnicity. Note: CDC life tables do not currently include NHPI or More than one race categories. Therefore, ranges are supplied based on applying life expectancy figures from the group with the lowest life expectancy (AI/AN) and the highest (Asian). Data for the More than one race category are not shown but are included in the figures for all races (bottom row).

Over 454,200 (32.9%) excess deaths occurred in people under age 65 (Table 1), accounting for ~14,209,200 (61.2%) YPLL; there were ~13,104,500 YPLL among persons ages 25-64 and ~1,104,700 YPLL among persons ages <25 (Table 1, Table S1-3). Raw counts, incidence rates, and observed-to-expected ratio for excess mortality varied by race/ethnicity among all age groups (Table 1, Figure 1, Figure S1-3). The highest rates of excess mortality and YPLL per capita were observed among the AI/AN (822 excess deaths per 100,000; ~405,700 YPLL) and Black populations (549 excess deaths per 100,000; ~4,289,200 YPLL, Table 1). The highest observed-to-expected ratios were documented among the AI/AN (1.34, 95% CI 1.31-1.37) and Hispanic (1.31, 95% CI 1.27-1.34) populations (Figure 1, Panel A). Greater YPLL per capita (and per excess death) among AI/AN, Black, Hispanic, and NHPI populations were observed (Figure 2, Table S4-5), reflecting younger mean/median ages of mortality (Figure S4-5), as compared with Asian and White populations.

**Figure 1.**
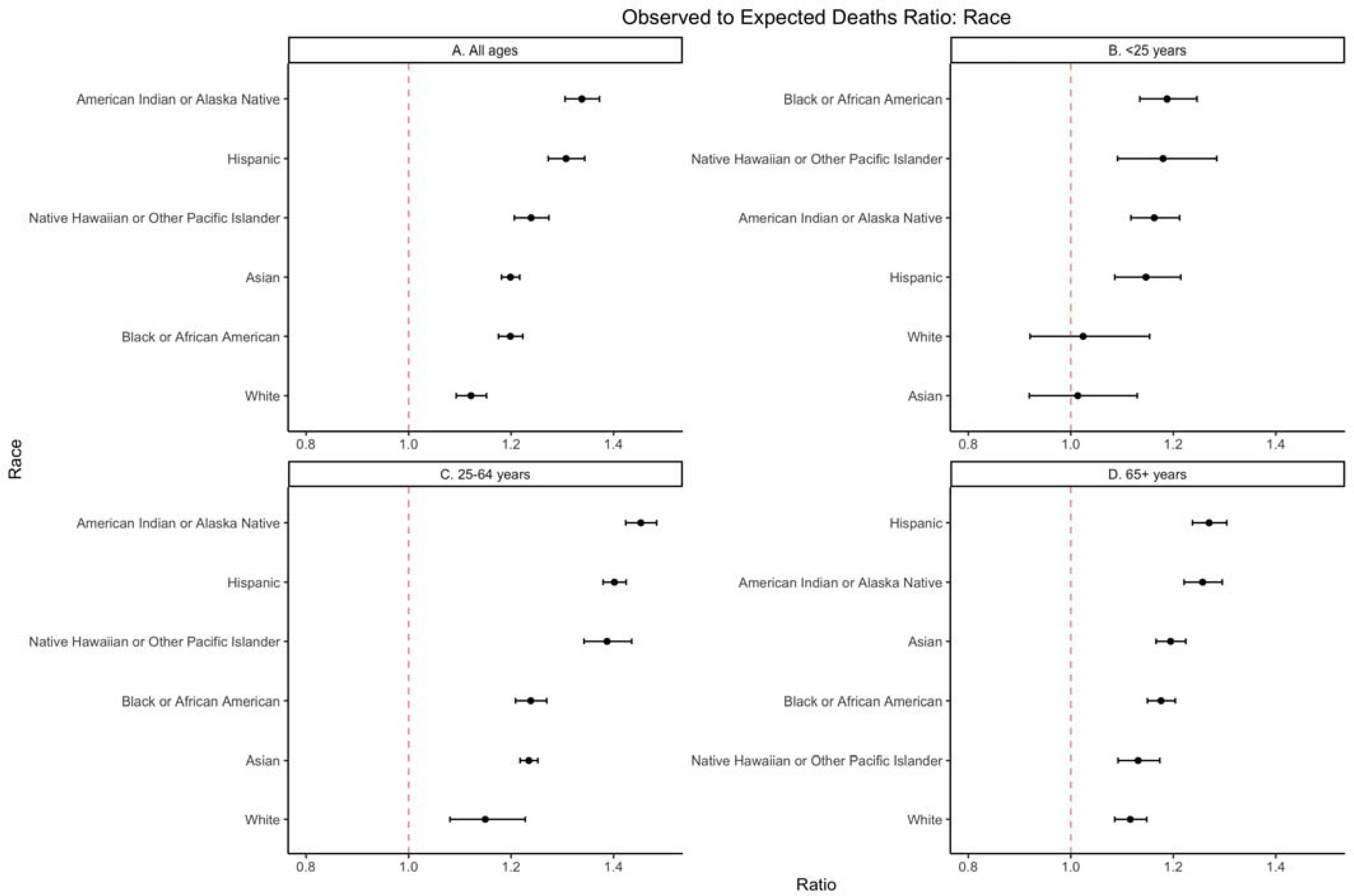
Observed-to-expected ratios for all-cause mortality during the pandemic period by race/ethnicity are shown from greatest (top) to least (bottom). For each group, the graphed ratio reflects raw observed deaths divided by the number of modeled expected deaths during the pandemic period for that specific population; 95% confidence intervals are shown as horizontal bars. The vertical line (1.0) means no change. A: All ages; B: ages <25 years; C: ages 25-64 years; D: ages ≥65 years.

**Figure 2.**
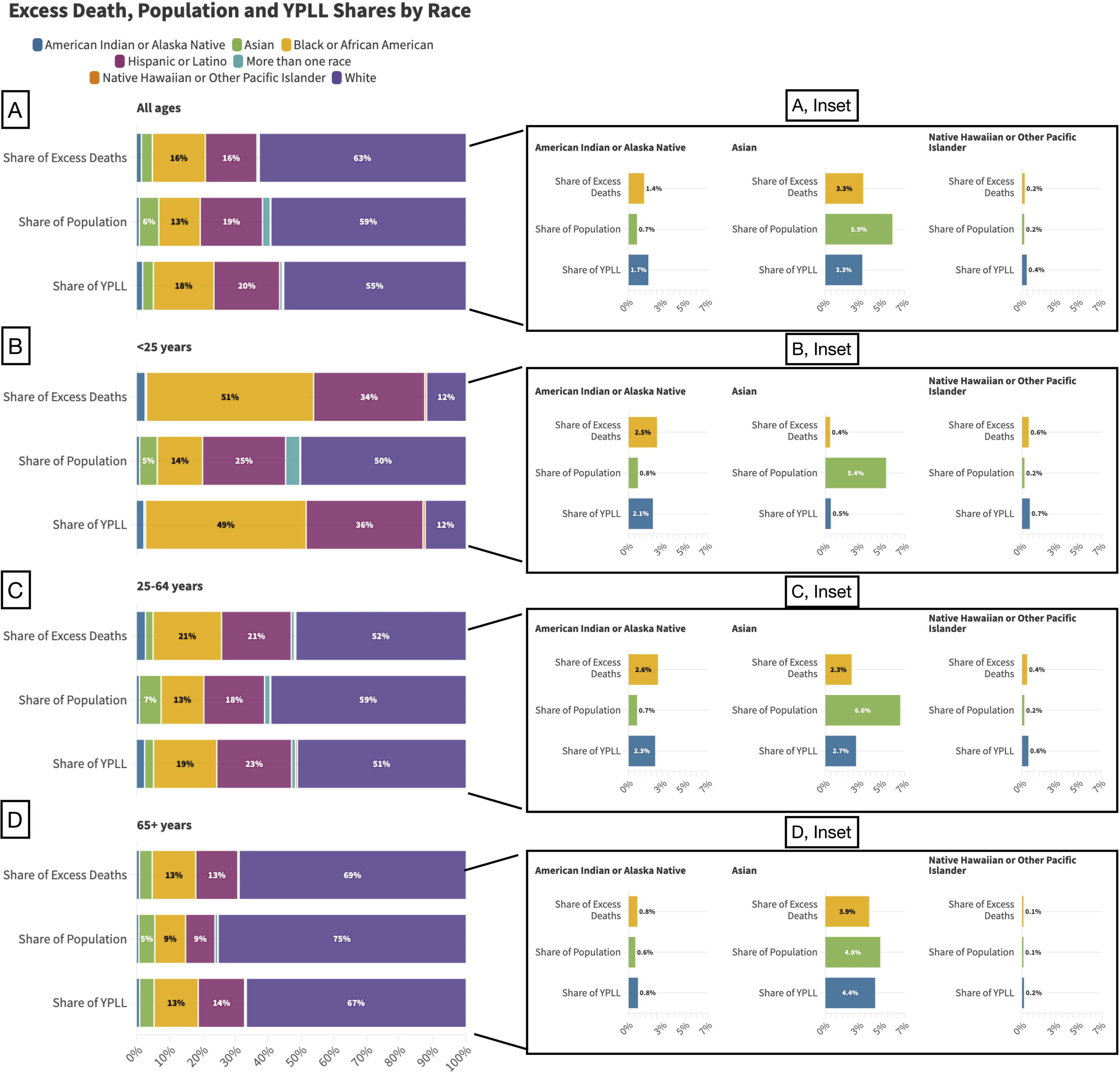
Shares of excess death, population and years of potential life lost (YPLL) by race/ethnicity and age (A: all ages; B: ages <25 years; C: ages 25-64 years; D: ages ≥65 years). In each panel, the middle bars show the population share for each race/ethnicity (“More than one race” group not analyzed elsewhere is included here for completion). Above the middle bar, the share of excess deaths by race/ethnicity is shown for the demographic. Below the middle bar, the share of YPLL deaths by race/ethnicity is shown for the demographic. Insets show demographics with smaller populations.

Among younger persons (<25 years), the number of excess deaths included ~9,000 Black (~542,90 YPLL), ~6,000 Hispanic (395,000 YPLL), ~400 AI/AN (~23,800 YPLL), and 100 NHPI people (~7,500 YPLL) (95% confidence intervals for excess mortality and corresponding YPLL among younger Asian and White people crossed 0) (Figure 1, Panel B, Table S1). The rates and observed-to-expected ratios are reported in Table 1 and Table S1.

Excess deaths among adults ages 25-64 years included ~213,300 White (~6,743,200 YPLL), ~97,400 Hispanic (~2,993,500 YPLL), ~97,300 Black (~2,534,600), ~11,500 Asian (~356,700 YPLL), ~11,700 AI/AN (~308,000 YPLL), and ~1,950 NHPI (~73,000 YPLL) people (Table S2). Incidence rates (Table S2) and observed-to-expected ratios were highest among AI/AN, Hispanic, and NHPI populations (Figure 1, Panel C). While older adults (≥65 years) accounted for 67% of excess mortality for all race/ethnicities combined, among AI/AN and NHPI populations, people ages <65 years accounted for most of the excess mortality (60.5% and 70.3%, respectively) (Table 1, Figure 2, Panels B-C, Table S1-2). The shares of excess mortality among those <65 years in each race/ethnicity were: NHPI (70.3%), AI/AN (60.5%), Black (46.6%), Hispanic (46.6%), White (25.2%), and Asian (24.1%) (Table 1).

For all race/ethnicity groups, the highest excess mortality incidence rates occurred among older adults (≥65 years, Figure 1, Panel D, Table S3), but among all age groups, the observed-to-expected ratios were lower for each population of older adults as compared to adults ages 25-64 (Figure 1, Panel D, Table 1, compare to Figure 1, Panel C and Table S2).

### Disparity rate ratios (DRR)

The share of excess mortality exceeded the share of the population among AI/AN (DRR; 1.4% share of excess deaths/0.7% population share=1.96), Black (DRR; 16.5% share of excess deaths/12.6% population share=1.31), and NHPI (DRR; 0.2% share of excess deaths=0.2% population share =1.12) populations (Table S4). Similar findings were documented by age group (Table S4), though among those ages <25 years, the Black population accounted for a majority (51.4%) of excess mortality despite representing only 13.8% of the age demographic population (DRR=3.72, Figure 2, Table S4).

### Covid and all-cause excess mortality

During the pre-vaccine era, the magnitude of excess mortality (per person-month) and between-group disparities were greater (Figure 3, Figure S4, Figure S6) than later in the pandemic. Monthly all-cause excess mortality correlated very strongly with Covid-19-specific mortality for the total population (Figure 3) and among adults ages 25-64 and >65 years (Figures S8-9). Weak correlations were observed between all-cause excess mortality and Covid-19-specific mortality for the population ages <25 years (Table S6; Figure S7). Cause-specific excess mortality and Covid-19 mortality demonstrated moderate or strong correlations according to several medical causes of death (Figure S7) and varied by race/ethnicity and age (Figures S10-13). Excess mortality for external manner deaths (e.g., accidents, homicide, unintentional overdose) was observed for all race/ethnicity groups. No significant correlations were detected between external manner deaths and Covid-19 mortality (Table S7).

**Figure 3.**
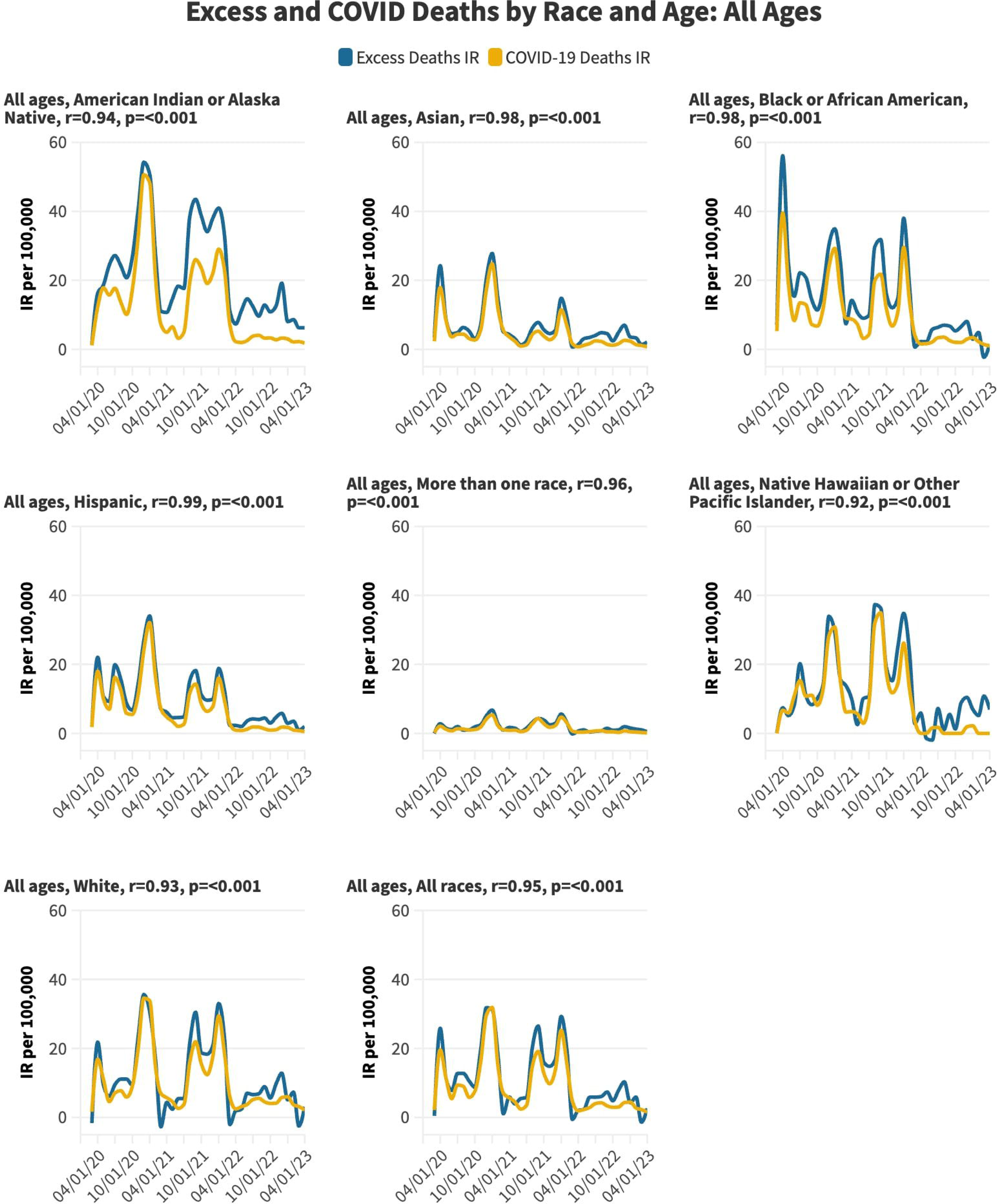
Monthly all-cause excess mortality and Covid-19-specific mortality by race/ethnicity. Blue lines show excess mortality; yellow lines show Covid-19-specific mortality. The Pearson correlation coefficient relating the lines are shown for each race/ethnicity, with their corresponding p values.

### Changes in all-cause mortality disparities during the pandemic

Established pre-pandemic disparities in all-cause mortality by race/ethnicity (age-adjusted RRs) increased during the pandemic (Figure 4, Panel A-C). Baseline all-cause mortality disparities were greatest among older adults, as were pandemic-associated increases (Figure 4, Panels D-L, Table S8).

**Figure 4.**
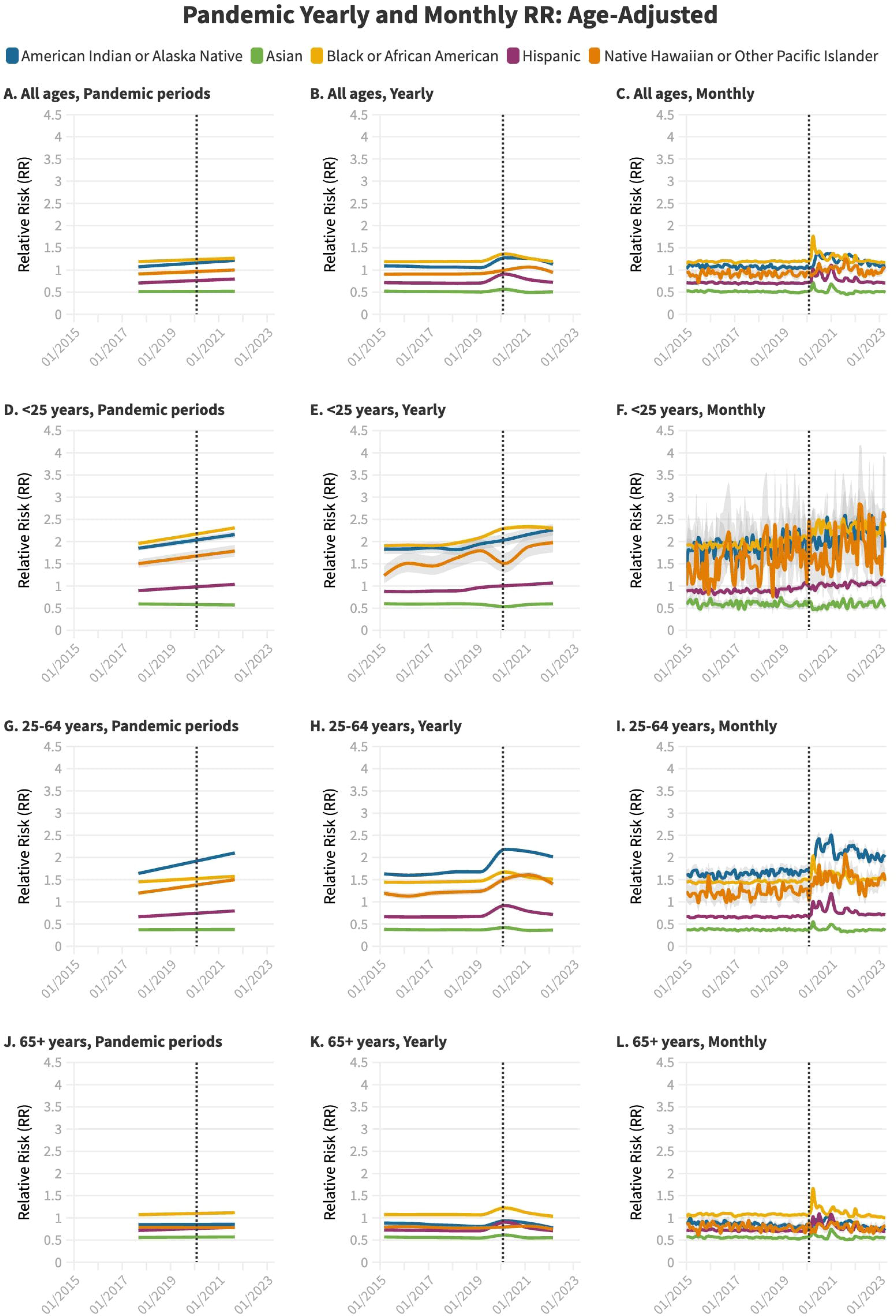
Pandemic yearly (left panels) and monthly (right panels) relative risk (RR) of age-adjusted all-cause mortality compared to the same-age White population (RR=1.0). For each age grouping (A-B: all ages; C-D: ages <25 years; E-F: ages 25-64 years; G-H: ages ≥65 years), yearly (left) and monthly RR are shown. AI/AN is shown in blue, Asian in green, Black in yellow, Hispanic in purple, NHPI in orange. 95% confidence intervals are shown for each line compared to the White population (note: some confidence intervals may be imperceptible due to narrow bandwidths). The vertical dashed line denotes the beginning of the Covid-19 pandemic period.

For the entire group, compared with the White population referent, AI/AN and Black populations experienced the greatest disparities during the pre-pandemic and pandemic phases. Among the AI/AN population, the pre-pandemic RR (1.07, 95% CI 1.06-1.08), increased to 1.22, 95% CI 1.21-1.22 (Table S8-13). Among the Black population, the pre-pandemic RR 1.19 (95% CI 1.19-1.19) increased to 1.26 (95% CI 1.26-1.27). Among the Asian, Hispanic, and NHPI groups (i.e., those with favorable pre-pandemic baselines relative to the White population, all-cause mortality RRs increased during the pandemic.

Among older adults, significant increases from baseline RRs occurred in only two groups: Black (1.07 pre-pandemic; 95% CI 1.07-1.07 to 1.11 pandemic; 95% CI 1.11-1.12) and Hispanic (0.72 pre-pandemic; 95% CI 0.71 - 0.72 to 0.79 pandemic; 95% CI 0.79 - 0.79) (Figure 4, Panels J-L). In the population ages 25-64, the AI/AN population experienced the highest pre-pandemic mortality disparity (1.64; 95% CI 1.62-1.66), and the largest RR increase (2.10; 95% CI 2.08-2.13) (Figure 4, Panels G-I, Table S8). Among those ages <25 years, the Black population had the highest mortality disparity at baseline (1.96; 95% CI 1.94-1.97) and the largest RR increase during the pandemic (to 2.31; 95% CI 2.29-2.33) (Figure 4, Panels D-F, Table S8).

Changes in pre-pandemic disparities were greater during Covid-19 waves (Figure 4). Changes in disparities peaked during the first or second pandemic year and then receded, but among the population ages <25, mortality RR continued to rise or remained stably elevated compared with pre-pandemic baselines (Figure 4, Table S11). By the third pandemic year, RRs for mortality had returned to pre-pandemic levels except for AI/AN and NHPI populations (Figure 4, Table S10-13). In analyses of the entire group, changes from pre-pandemic baselines were mostly abolished during the vaccine era. (Table S9).

## Discussion

This study documents massive mortality disparities during the US Covid-19 public health emergency, accounting for 252,000 more excess deaths and 5.2 million more YPLL than would have occurred without the disparities. Though all racial/ethnic groups experienced pandemic-associated excess mortality, AI/AN people experienced the highest rate of all-cause excess mortality and the greatest disparity; their share of excess mortality was nearly twice what would have been projected, based on population shares. Age-stratified analyses revealed important disparities in the <65 population overall, with the largest increases in mortality (observed-to-expected ratio) occurring among adults ages 25-64 years in each race/ethnicity. In the population ages <25 years, Black people comprised a majority (51%) of all-cause excess deaths despite representing 14% of the demographic population. Excess mortality among <25 years) was documented among AI/AN, Hispanic, and NHPI but not among the Asian or White populations (Figure 1). Additionally, a long-standing (pre-pandemic) finding of lower all-cause mortality rate in the Hispanic population compared with White and Asian population was narrowed and even reversed during Covid-19 waves.^15^

This analysis extends the existing literature in several ways. First, we utilized age-stratified component excess mortality modeling, rather than age-adjustment, which identified substantial differences in excess mortality by race/ethnicity, particularly among the population <65 years; this method allowed us to more accurately characterize ground conditions, permitted YPLL estimates within groups, and unveiled profound, long-term implications related to excess mortality that was concentrated among the younger demographics. Second, these data offer detailed accounts of disparities which spanned the entire pandemic. Despite early awareness of disparities, some persisted—albeit disparities decreased in older populations after vaccines became available (and after the initial Omicron wave) (Figures S1-3, Tables S8-S13). Third, we used pre-pandemic and pandemic age-adjusted, all-cause mortality rates to establish the finding that existing mortality disparities among certain groups widened during the Covid-19 era, rather than merely having been amplified by global increases in incident rates, an effect especially pronounced among working-aged adults. Known Covid-19 waves were associated with sudden changes in disparities.^16^ Fourth, we conducted exploratory analyses that identified increases in non-Covid-19 causes of mortality during the pandemic; increases in select medical causes of death correlated with Covid-19 mortality waves, which implied that many “non-Covid” deaths were directly or indirectly caused by the virus or secondary effects. The result is the most comprehensive assessment of US Covid-19 pandemic-associated mortality disparities by race/ethnicity and age.

Importantly, we demonstrate that the pandemic exacerbated historical disparities due to strata in social determinants of health, structural inequality, and racism. Compounding the inherent value of the loss of life/life-years, associated economic productivity losses resulting from identified excess mortality in working-aged and younger adults have important implications, as health and economic affluence are correlated.^25^ Meanwhile, less dramatic disparities among older populations, before and during the pandemic, likely reflect “healthy survivor” effects among groups with higher death rates in younger demographics.

Given that race and ethnicity are social constructs, the magnitude of these findings cannot be explained by genetic differences. Possible biological and sociological explanations for our findings include higher rates of pre-existing conditions associated with poor Covid-19 outcomes, higher probability of viral exposure during the pre-vaccine era (reflecting higher shares of essential workers in certain populations, initial lack of equitable access to Covid-19 testing, decreased remote learning or access to school-implemented mitigation measures, and distrust in public messaging focused on infection prevention), decreased access to healthcare (including *de facto* segregated systems), and racial biases in Covid-19 treatment^17–22^—all manifestations of inequities stemming from structural racism and poverty disparities. Initial skepticism of Covid-19 vaccines related to historical medical mistrust was likely another factor; during the early vaccine rollout, receipt of vaccinations was lower among Black people^23^; however, decreased disparities in all-cause mortality in Black people coincided with increased vaccine uptake later in 2021, suggesting successful messaging from trusted leaders.^24^

To prepare for future pandemics, efforts to protect high-risk groups—utilizing evidence-based policy, equitable distribution of resources, and improving infrastructure—are essential. To achieve this level of preparation, systemic factors must be addressed. In addition to preparation efforts, just-in-time responses should be directed toward high-risk communities during emergencies (pandemics, natural, or human-caused disasters).^26^ For example, policies providing appropriate and cost-free isolation for essential workers would have prevented chains of Covid-19 transmission to higher-risk people who resided in multi-generation households, especially during pre-vaccine and pre-therapeutic eras.^27–29^ Similarly, efforts to increase vaccination uptake and implement effective therapeutics among at-risk populations would have reduced disparities.^23,30–36^ For the overall population and among all adults (>25 years), existing race/ethnicity mortality disparities widened at the onset of the pandemic and during Covid-19 waves but narrowed during the vaccine era. The successes of mass vaccination efforts in 2021 mitigated access barriers but were not replicated during subsequent booster campaigns.

Differences in vaccination rates and booster receipt in at-risk persons may partly explain why the AI/AN population experienced higher excess mortality later in the pandemic, as this population predominantly lives in underserved areas where access barriers are more common.^21,22^ Lastly, the pandemic was associated with increases in deaths by external manner (e.g., homicide, unintentional overdose, albeit not suicide), especially in younger age groups, with marked between-group differences.^37,38^ Unlike several medical causes of death, external manner deaths were not temporally correlated with Covid-19-mortality (Figures S10-13).^39^

This study has limitations. First, calculating all-cause excess mortality relies on estimating expected deaths. While pre-pandemic trends would likely have continued without the pandemic, certainty is not possible. Second, the determination/coding of race/ethnicity at the time of death is not standardized, which could result in inaccuracies. However, these issues are unlikely to have changed during the pandemic. Third, the exploratory cause/manner of death analyses are subject to death certificate inaccuracies. Fourth, instead of granularity, we defined three large age groups to capture generational differences, which may have had small effects on the final estimates.

This study describes substantial mortality disparities by race/ethnicity in the US during the Covid-19 public health emergency period, particularly among younger populations, often dramatically exceeding pre-pandemic disparities. While pandemics are inevitable, disparities are not. The need to address the conditions that create health disparities—before the next public health crisis—is evident.

## Data Availability

All data analyzed in the present study are publicly available.

