## Supplemental Appendix for "Age-Specific All-Cause Mortality Disparities by Race and Ethnicity During the COVID-19 Pandemic"

**Table of Contents**

Supplemental methods (with Methods Table).

Tables S1-S3: Expected, observed and excess deaths by race/ethnicity; Ages <25 years (S1), Ages 25-64 years (S2), Ages ≥65 years (S3).

Figure S1: Cumulative excess mortality per 100,000 persons by race/ethnicity and age group.

Figure S2: Cumulative excess mortality (raw) by race/ethnicity and age group.

Figure S3: Monthly excess mortality per 100,000 persons by race/ethnicity and age group.

Table S4: Disparity rate ratio between share of excess deaths and share of population, by race/ethnicity and age group.

Table S5: Years of potential life lost by race/ethnicity and age group.

Figure S4: Share of years of potential life lost by race/ethnicity and age group.

Figure S5: Years of potential life lost by race/ethnicity and age group.

Figure S6: Excess mortality per 100,000 persons by race/ethnicity and vaccine period, ages 25-64 years and ≥65 years.

Figures S7-9: Covid-19-specific and all-cause excess mortality per 100,000 persons by race/ethnicity; Ages <25 years (S7), Ages 25-64 years (S8), Ages ≥65 years (S9).

Table S6: Pearson correlation between all-cause excess mortality and Covid-19-specific mortality by race/ethnicity and age group.

Figures S10-13: Changes in cause-specific mortality and correlation to Covid-19-specific mortality. Observed deaths per 100,000 persons by race/ethnicity, age group and UCD – ICD Chapter: All ages (S10), Ages <25 years (S11), Ages 25-64 years (S12) and Ages ≥65 years (S13).

Table S7: Correlation between cause of death and COVID deaths, by race and age group.

Table S8: Relative risks by age and race/ethnicity, pre-pandemic and pandemic periods.

Table S9: Relative risks by age and race/ethnicity by vaccine period.

Tables S10-13: Relative risks by age and race/ethnicity, by pandemic year, All ages (S10) Ages <25 years (S11), Ages 25-64 years (S12) and Ages ≥65 years (S13).

**Supplemental Methods.**

**Excess Mortality**

Population estimates: Projected yearly changes were divided by 12 and applied to each calendar month to achieve smooth changes at the start of each year. The smoothed projected monthly populations for each group were applied to the mortality sARIMA model as a covariate to overcome stationarity because the number of expected deaths is dynamically influenced by the number of people in each group at any time. Monthly projections enabled the capture of seasonal trends.

We computed monthly and cumulative excess mortality incidence rates (per 100,000, using mean pandemic population for each group), and observed-to-expected mortality ratios were determined for all available age groups and race/ethnicity groups. A growing/rolling study period was created for each successive month during the 38-month study period for cumulative excess mortality. For example, March of 2020 was a study period, March-April of 2020 was a study period, meaning that the 38th study period (March 2020-April 2023) encompassed the entire study period. In addition, a comparison of excess mortality was made between the pre-vaccine period and the vaccine period (the vaccine period was measured from March 2021 on for persons ages ≥65 years, and May 2021 for persons ages 25-64).

The monthly 95% confidence interval [CI] boundaries were derived directly from the sARIMA model using the auto.arima function in the R statistical software. The cumulative period CI boundaries were obtained through the 5,000 simulation samples from the estimated sARIMA model for each age group (i.e., for each of the 38 study periods, 95% CIs were separately bootstrapped for each age group within each race/ethnicity and the total all-age composites.

Further, due to the effect of the pandemic on the size of the population, we adjusted the monthly populations to account for cumulative excess mortality during the public health emergency. For 12 months within all years, we defined the projected population for three age groups within each race/ethnicity group and age group (ages <25, 25-64 and ≥65 years), (denoted by $k$, where $1\leq k\leq4$) in month $i$ of 2020 was denoted as $n_{i,k}$; the original point estimate for all-cause deaths was denoted as $\hat{m}_{i,k}$; the sum of all previous months’ excess death were denoted as $P_{i-1,k}$ (starting from March 2020). Therefore, the corrected expected deaths for month $i$ (from March 2020) after correction ($\hat{m}_{i,k\_adj}$) satisfies:

$$\hat{m}_{i,k\_adj}=\left( n_{i,k}-P_{i-1,k} \right)\times\frac{\hat{m}_{i,k}}{n_{i,k}}$$

The above equation also holds for the study period months in 2021-2023, where $P_{i-1,k}$ denotes the sum of all previous months’ excess death.

**Model validation**

To test accuracy, sARIMA models were trained on monthly mortality data and population estimates as described (2012-2016 baseline) to project expected deaths from March 2017-February 2020. Because single-race data are not available for these years, the module was tested on alternate data (a 72-component model of the US assembled from 9 Census Divisions, 4 age groups, and 2 genders). In the 3-year validation period, the observed to expected ratio was >0.99 (8,518,962 modeled expected deaths, 8,575,330 observed deaths); 50% of monthly expected values were within ±3% of the unblinded observed values, 89% of monthly expected values were within ±5%, and 100% of monthly expected values were within ±10%. Yearly and total performance are shown below.

**Methods Table**

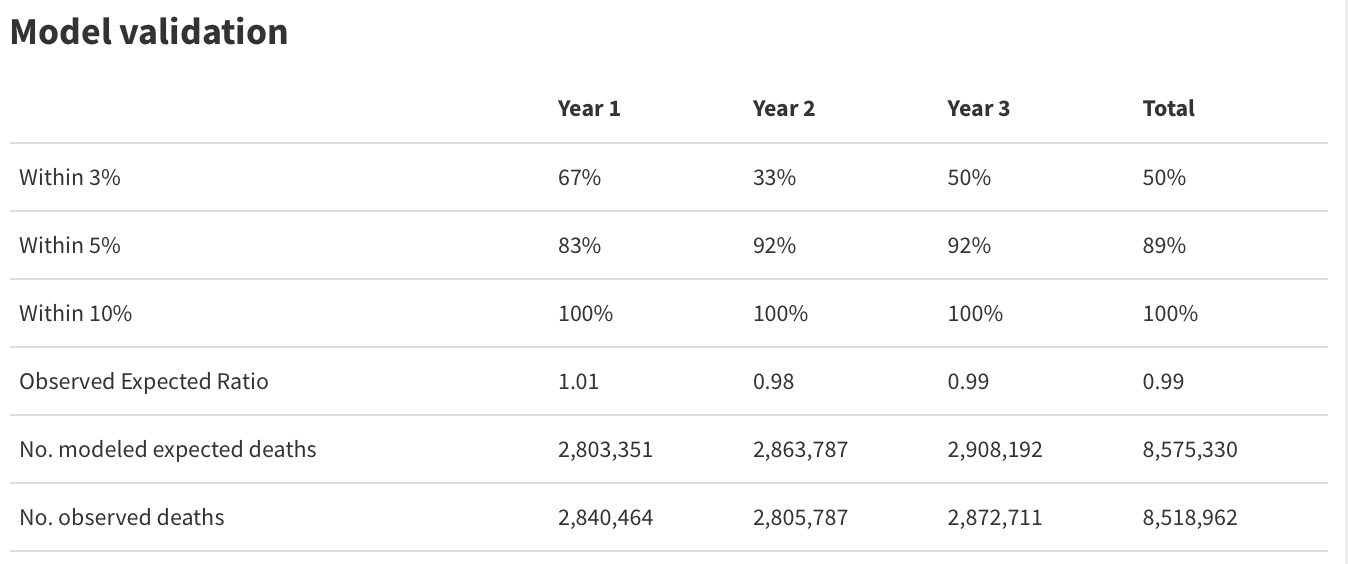

**Potential Excess Deaths Averted.**

The number of deaths averted was calculated by subtracting the observed-to-expected all-cause mortality ratio in the same-age White population from the observed-to-expected ratio for each individual population and multiplying the difference by modeled expected deaths that group (a second comparison using the same-age Asian population was also done). See: *Renton, B., Du, C., Chen, A.J. et al. State-Level Excess Mortality and Potential Deaths Averted in US Adults During the Delta and Omicron Waves of COVID-19. J GEN INTERN MED (2023). https://doi.org/10.1007/s11606-023-08374-2*

**Years of Potential Life Lost (YPLL) and Potential YPLL Averted.**

We assumed that within a 10-year age group, the distributions of excess deaths for each single year were like those seen among observed deaths during the pandemic period. Because no tables are available for NHPI or More than one race groups, we provided a range using the greatest (Asian) and least (AI/AN) life table values found among the other groups. For excess deaths averted and YPLL averted, the life expectancy estimates from Asian population were used.

Further, to determine the average YPLL per excess death, we divided the calculated YPLL by modeled excess deaths for each demographic.

The share of excess deaths and years of potential life lost from each of the three large age groups was determined for each race/ethnicity and age group by dividing the number of excess deaths in each age group and dividing by the total excess deaths for that race/ethnicity. For the share calculations (but not elsewhere), negative excess mortality was considered as 0.

Potential YPLL averted was calculated by applying the number of excess deaths averted (as above) to life expectancy tables (as above).

**Disparity Rate Ratios**

Disparity rate ratios (DRR) were calculated by dividing share of excess mortality in a demographic by the corresponding population share of that demographic. For example, for the all-ages analysis, AI/AN people represented 1.4% of excess mortality, which comprising slightly under 0.7% of the overall population. Therefore, the DRR=1.4%/0.7=1.96 with 95% confidence intervals (1.82-2.11) reflecting the corresponding excess mortality bandwidth.

**Cause-Specific Excess Mortality**

The cause-specific excess mortality module was generated using a similar approach as the all-cause excess mortality model, with two exceptions due to data limitations. First, we did not correct for the lower-than-expected population during the pandemic, owing to cumulative excess mortality; Second, the pre-pandemic baseline period was only available for 2018-2020, due to changes in the CDC’s method of reporting mortality data (i.e., the transition from bridged to single races/ethnicity).

We modeled cause-specific expected monthly deaths for each race/ethnicity and age group during the study period according to the International Classification of Diseases, Tenth Revision (ICD-10), limiting our analysis for each race/ethnicity and age group to causes for which the average monthly deaths were >25 during the pre-pandemic period, to avoid modeling causes of death with suppressed data (CDC does not report monthly causes of death whose counts are 1-9 deaths). A heat map was produced from the excess death incidence rate for each month. Total excess mortality from each modeled cause was determined (per 100,000 people).

**Table S1.** Expected, observed and excess deaths by race/ethnicity, ages <25 years.

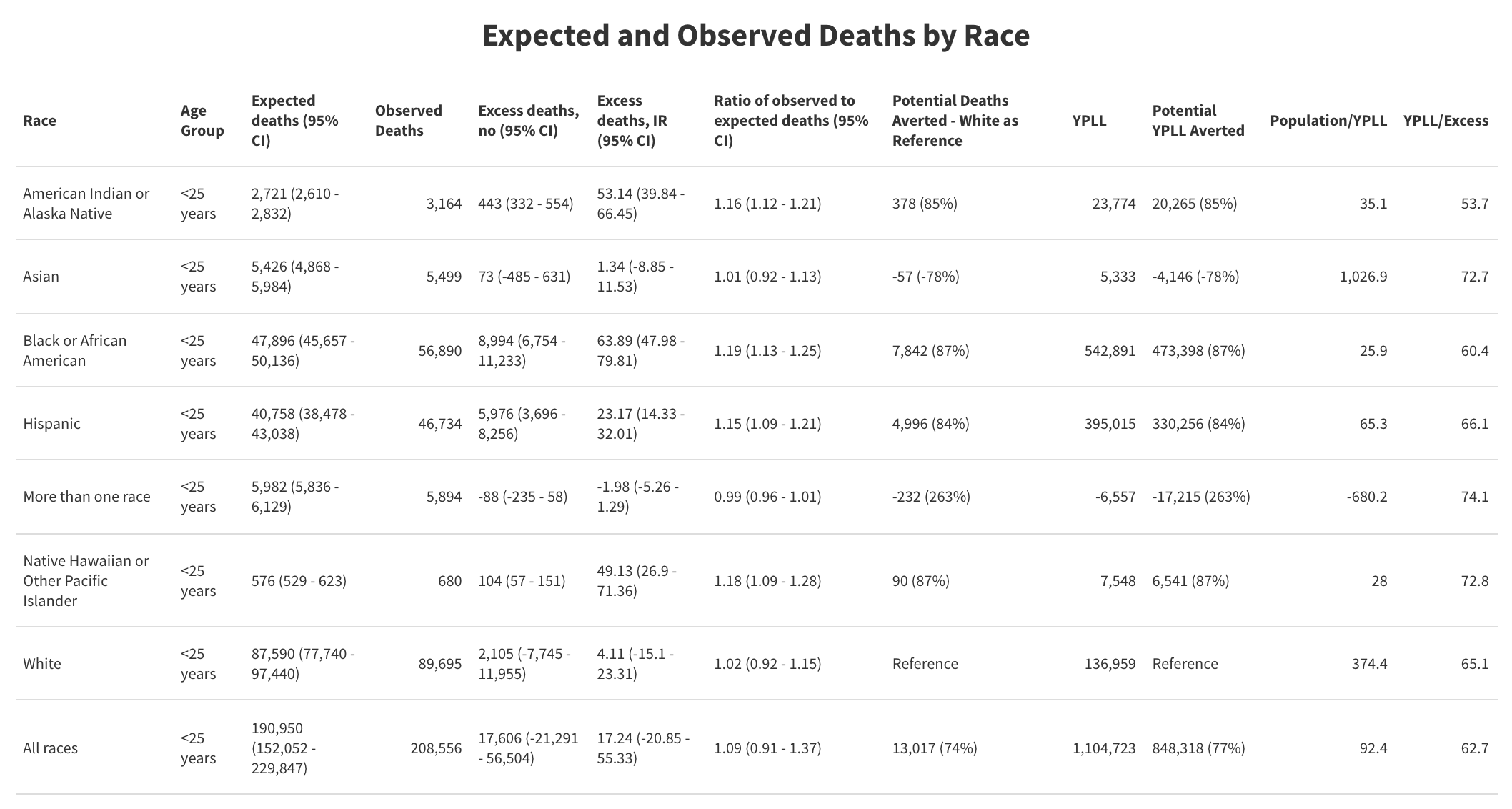

**Table S2.** Expected, observed and excess deaths by race/ethnicity, ages 25-64 years.

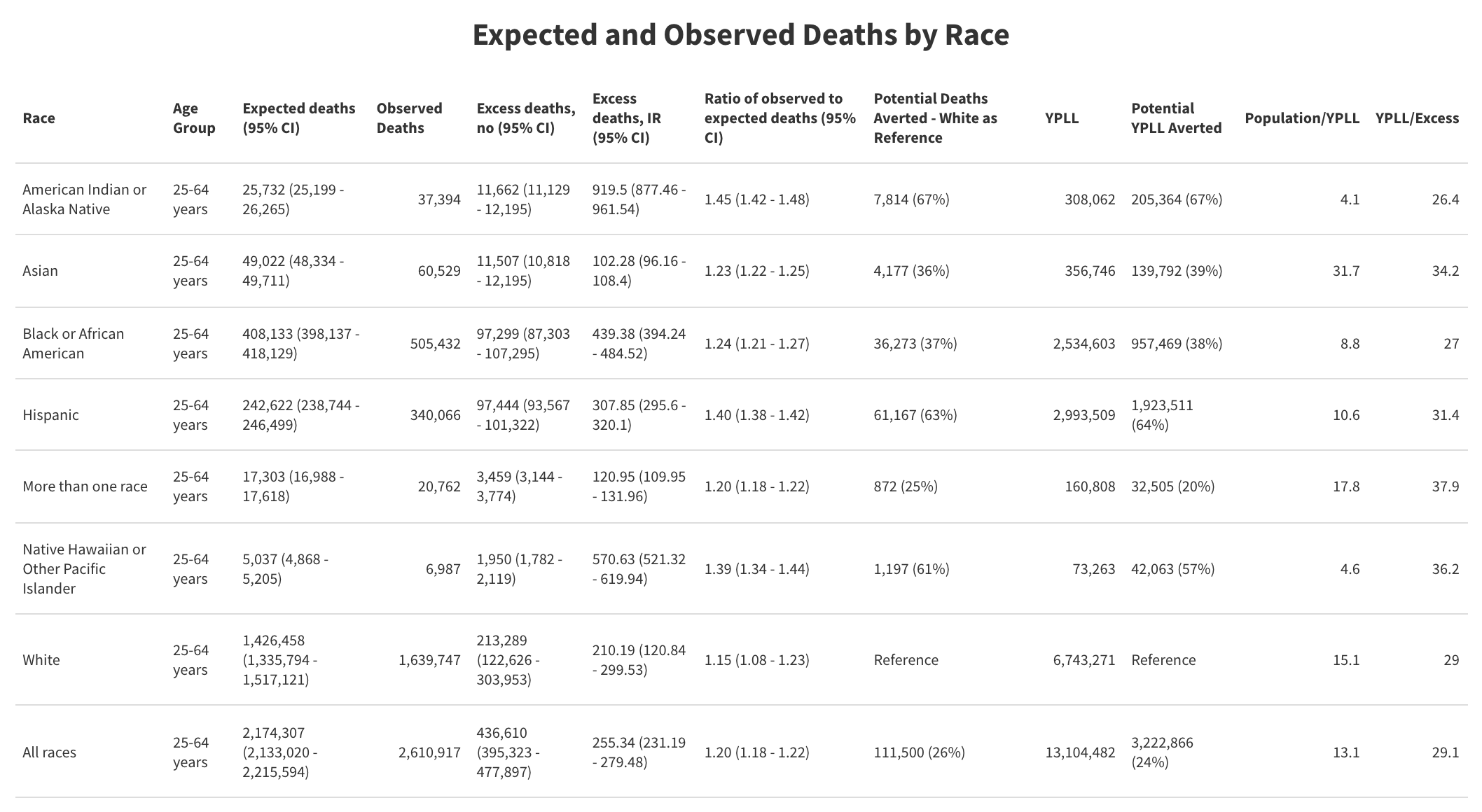

**Table S3.** Expected, observed and excess deaths by race/ethnicity, ages ≥65 years.

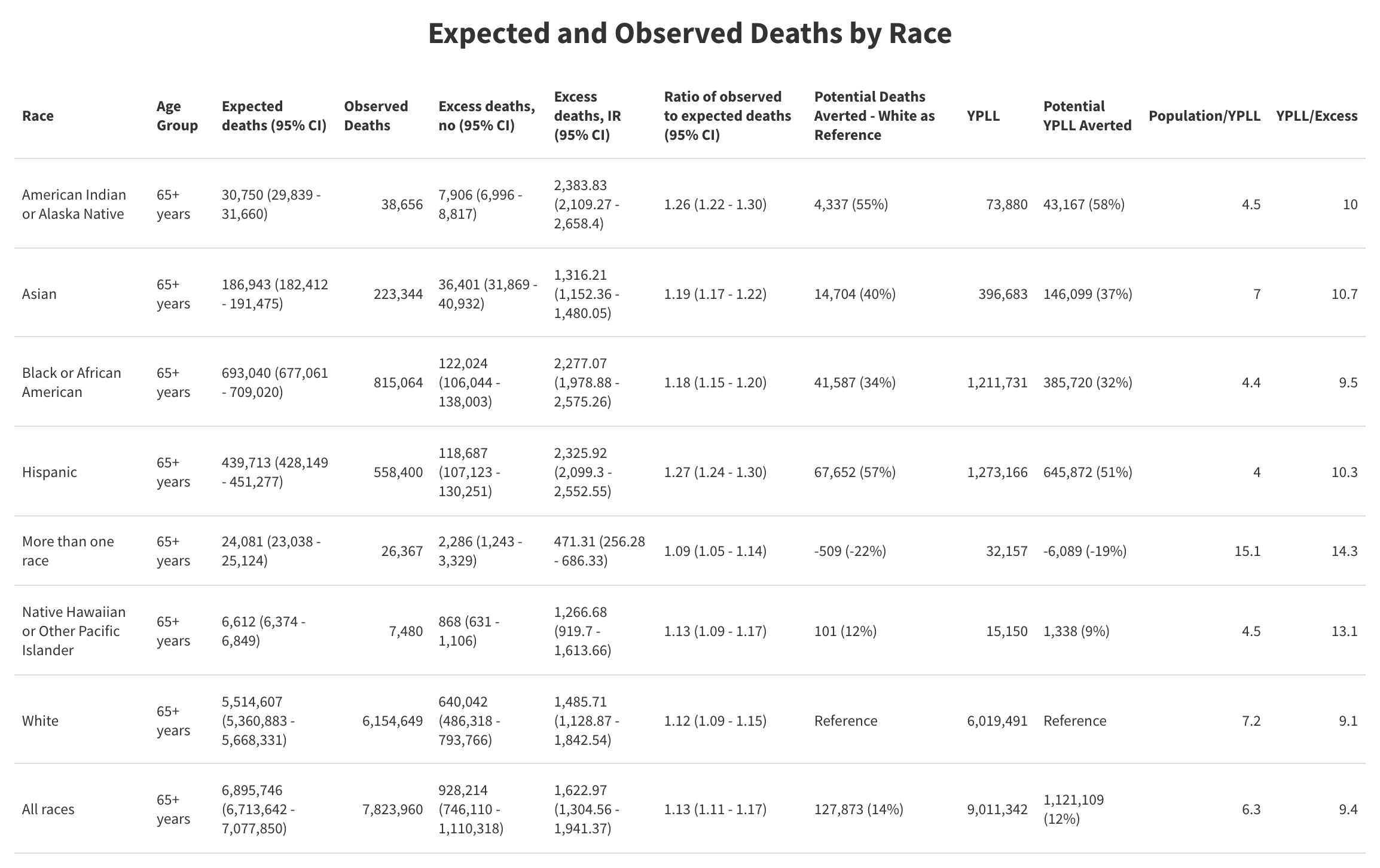

**Figure S1.** Cumulative excess mortality per 100,000 persons by race/ethnicity and age group. 95% confidence intervals are shown for all categories but may not be visible due to narrow bandwidths.

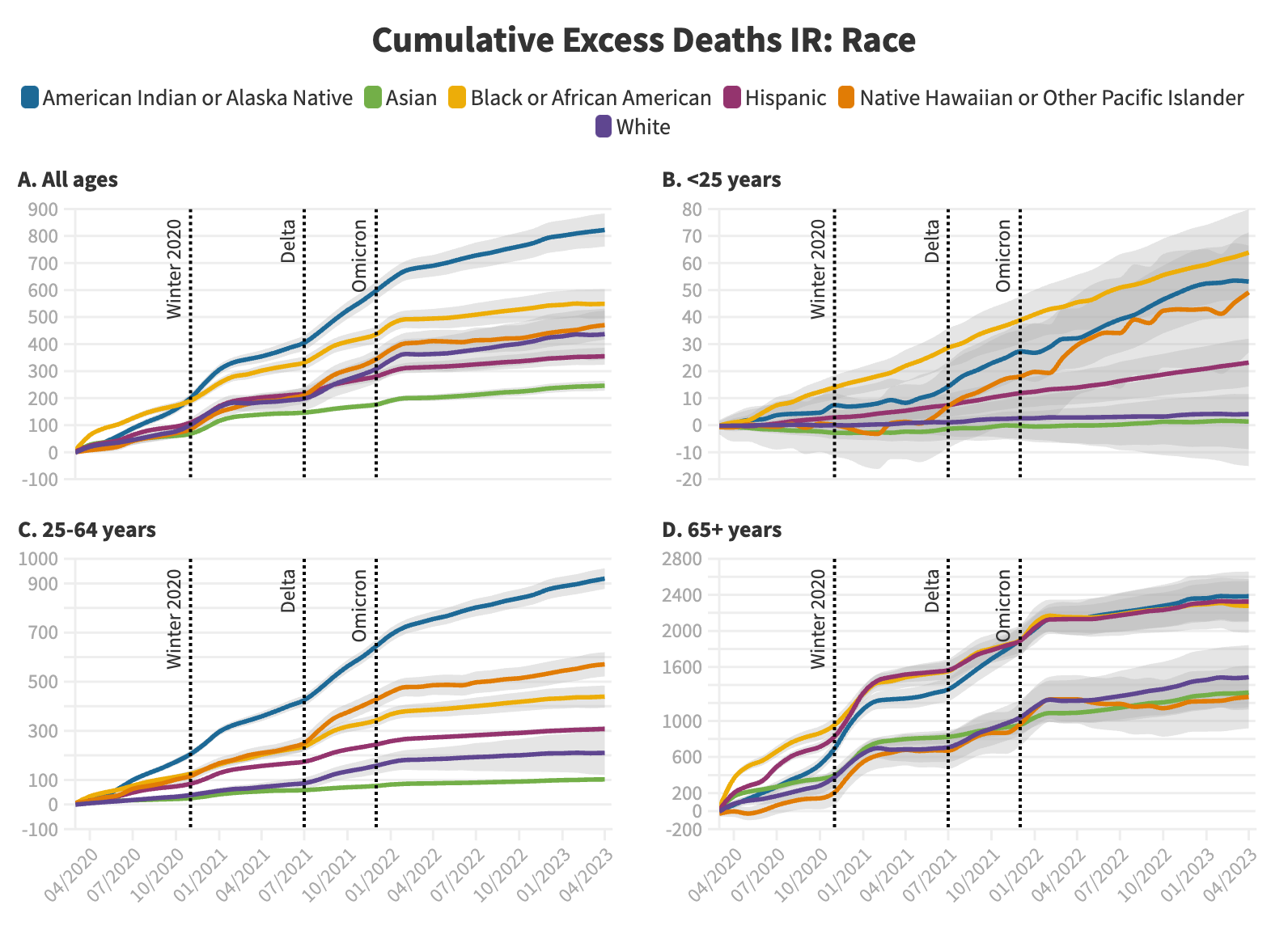

**Figure S2.** Cumulative excess mortality (raw) by race/ethnicity and age group. 95% confidence intervals are shown for all categories but may not be visible due to narrow bandwidths.

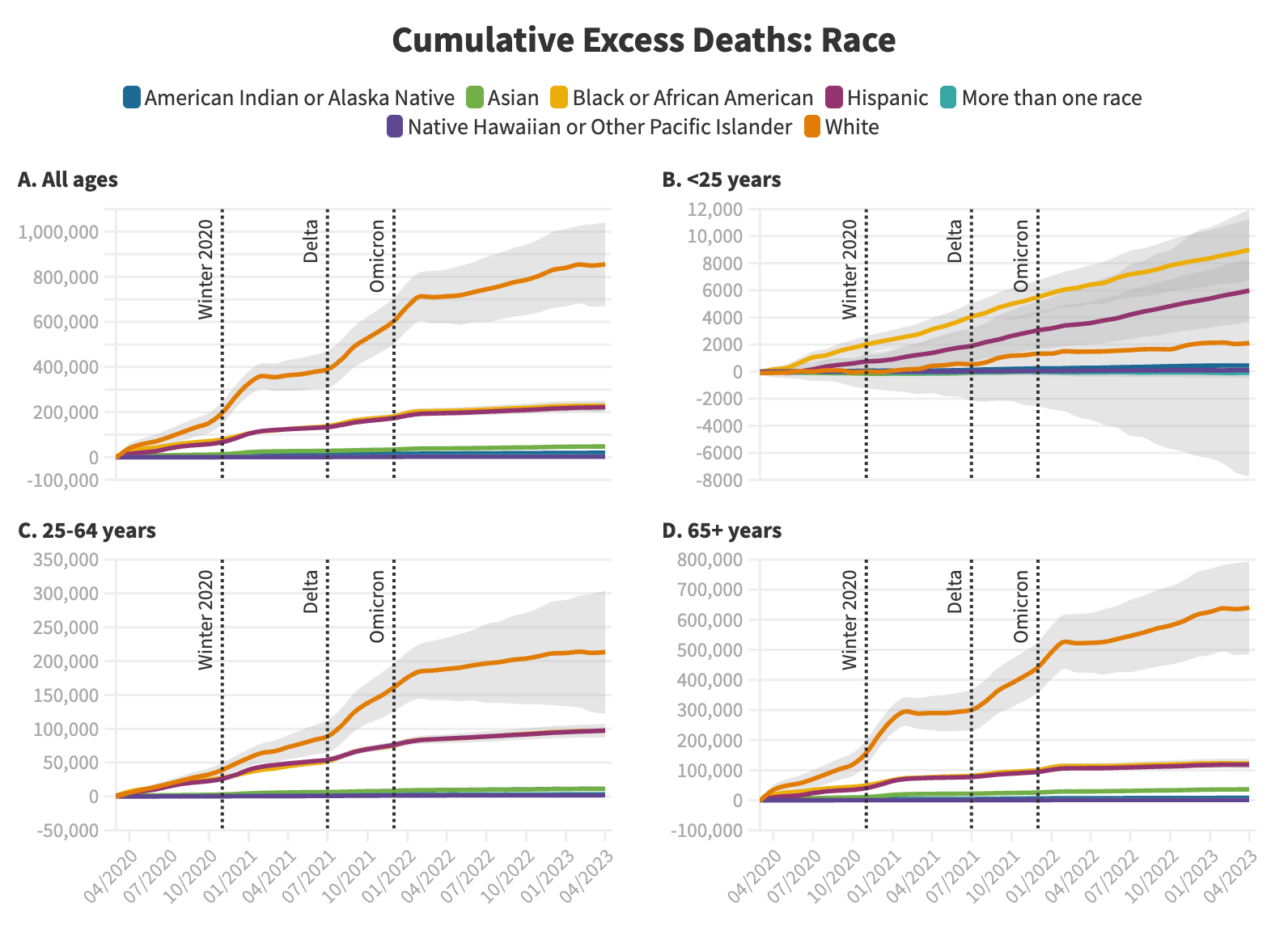

**Figure S3.** Monthly excess mortality per 100,000 persons by race/ethnicity and age group. 95% confidence intervals are not shown (for clarity).

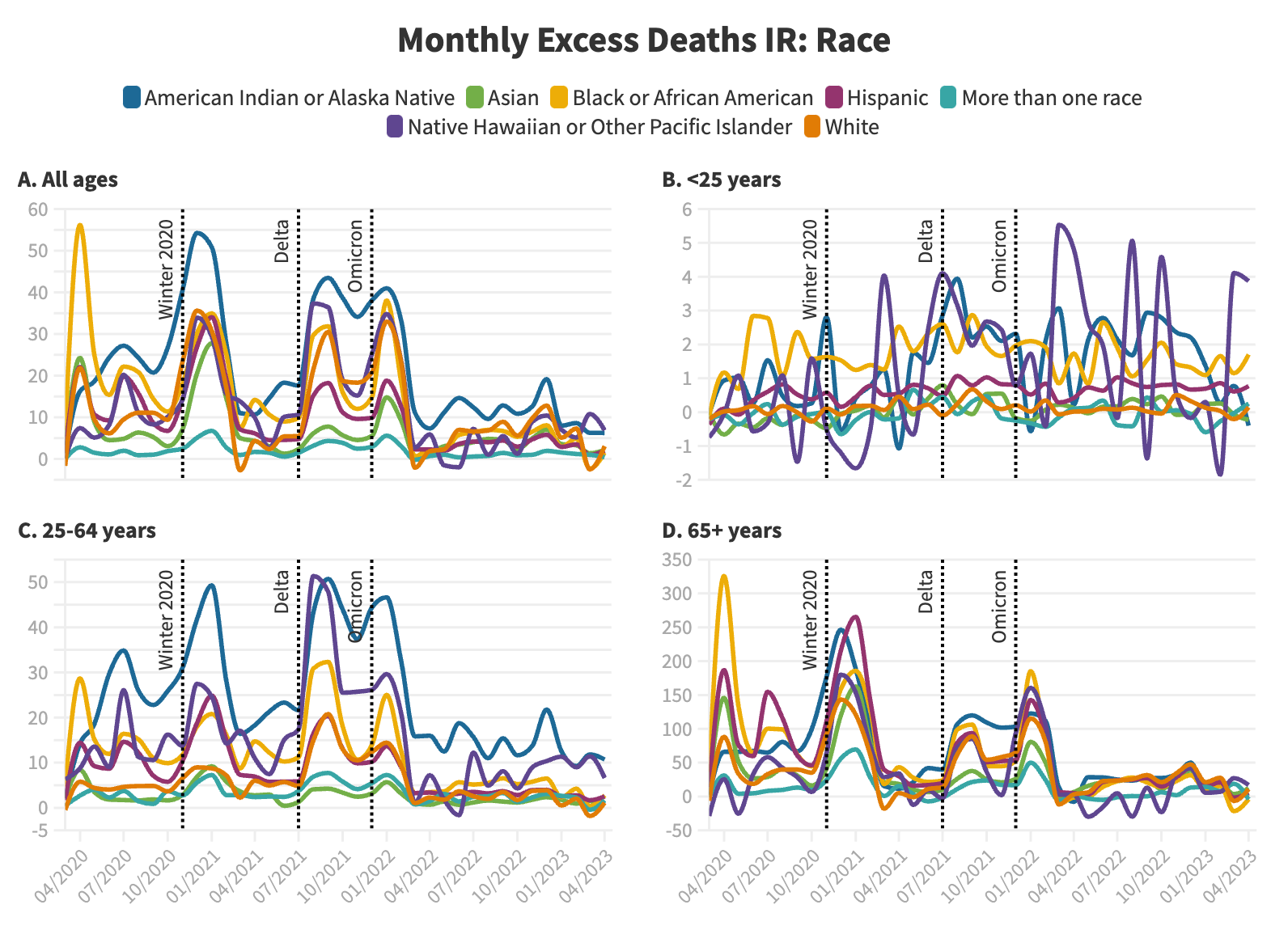

**Table S4.** Disparity rate ratio between share of excess deaths and share of population, by race/ethnicity and age group.

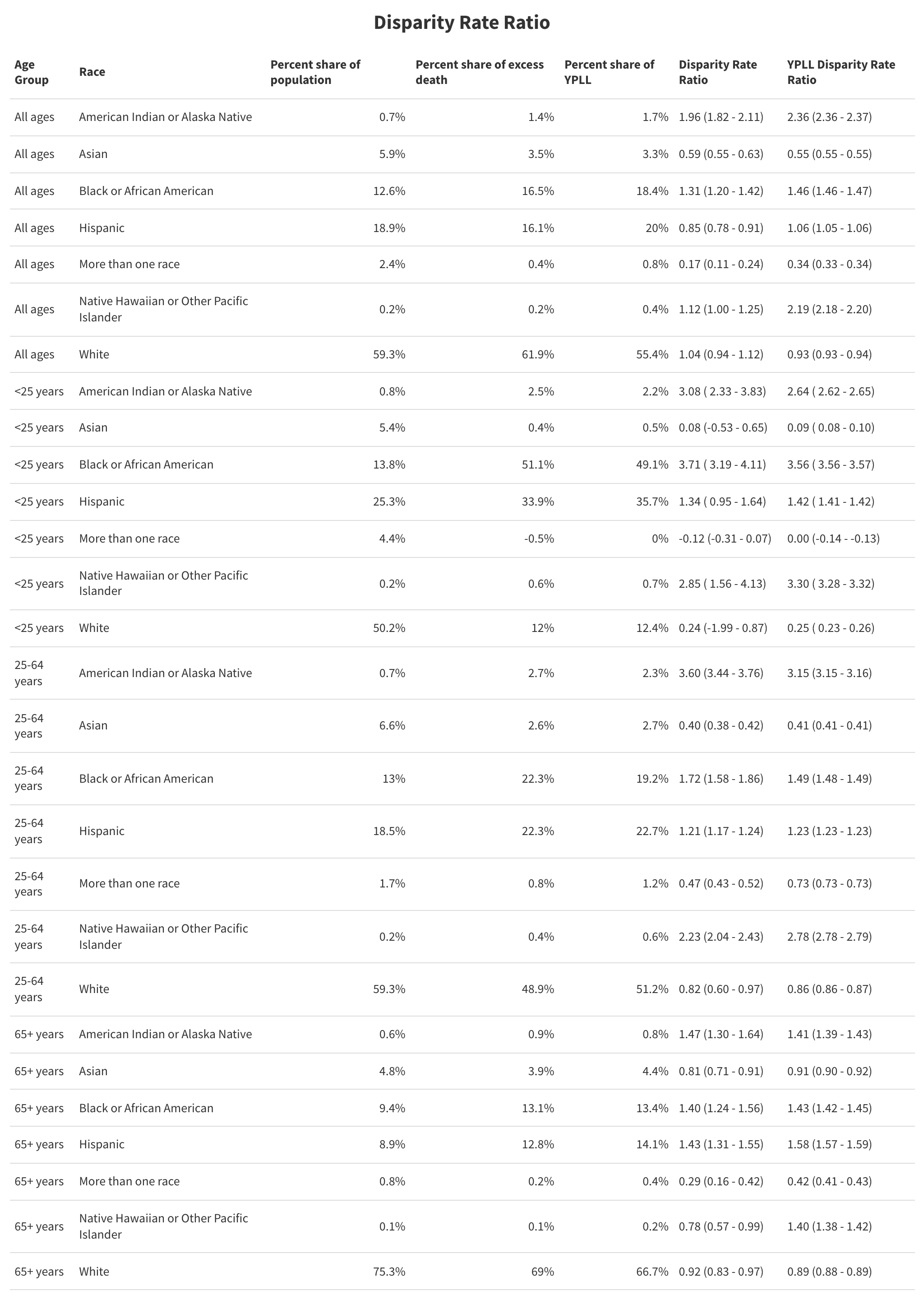

**Table S5.** Years of potential life lost (YPLL) by race/ethnicity and age group. For NHPI and More than one race categories, YPLL were determined using life expectancy for the Asian category, because CDC life tables do not include estimates for these populations.

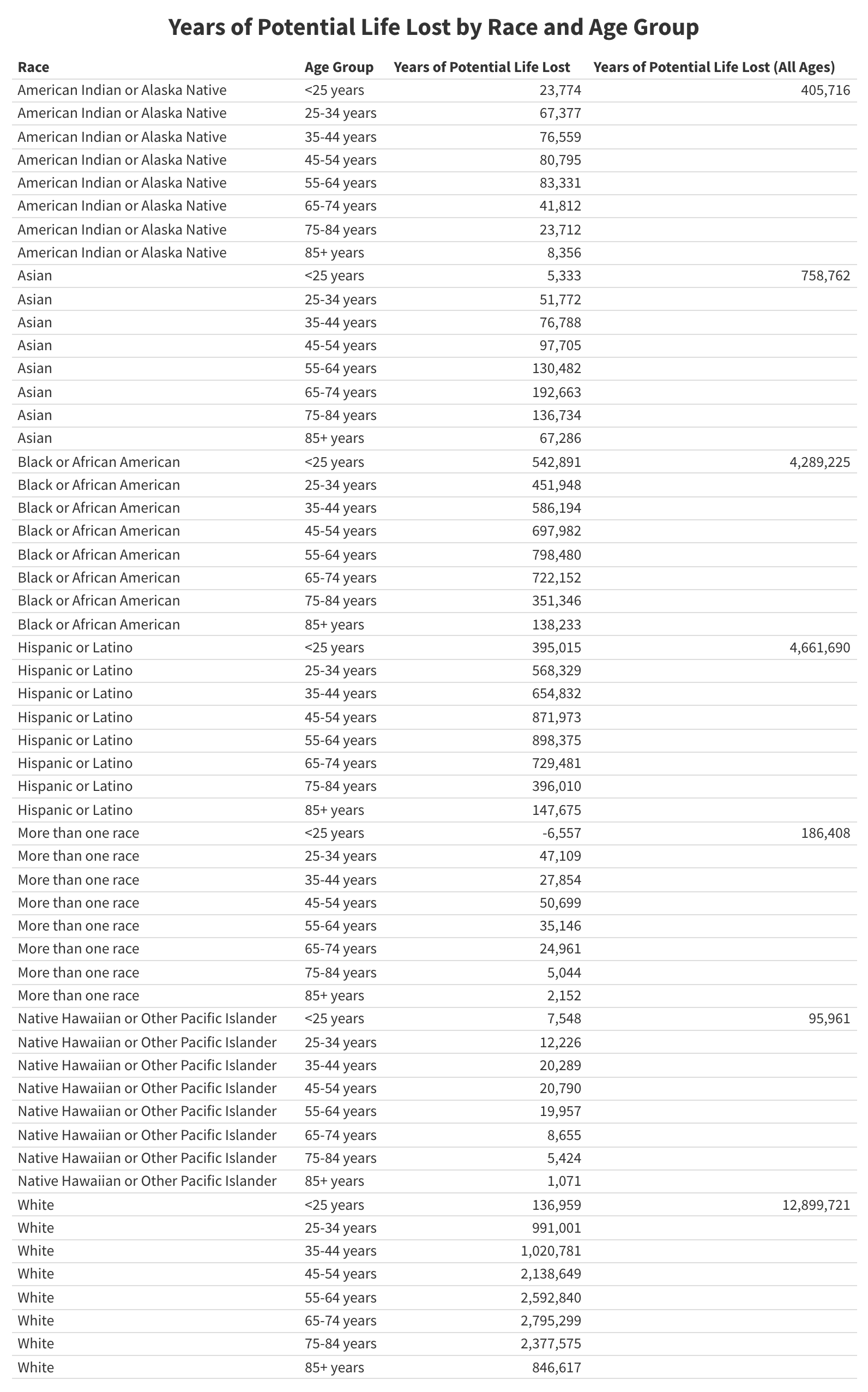

**Figure S4.** Share of years of potential life lost by race/ethnicity and age group (10-year groupings with the exception of <25 and ≥85 years, which were measured as one group to avoid suppression).

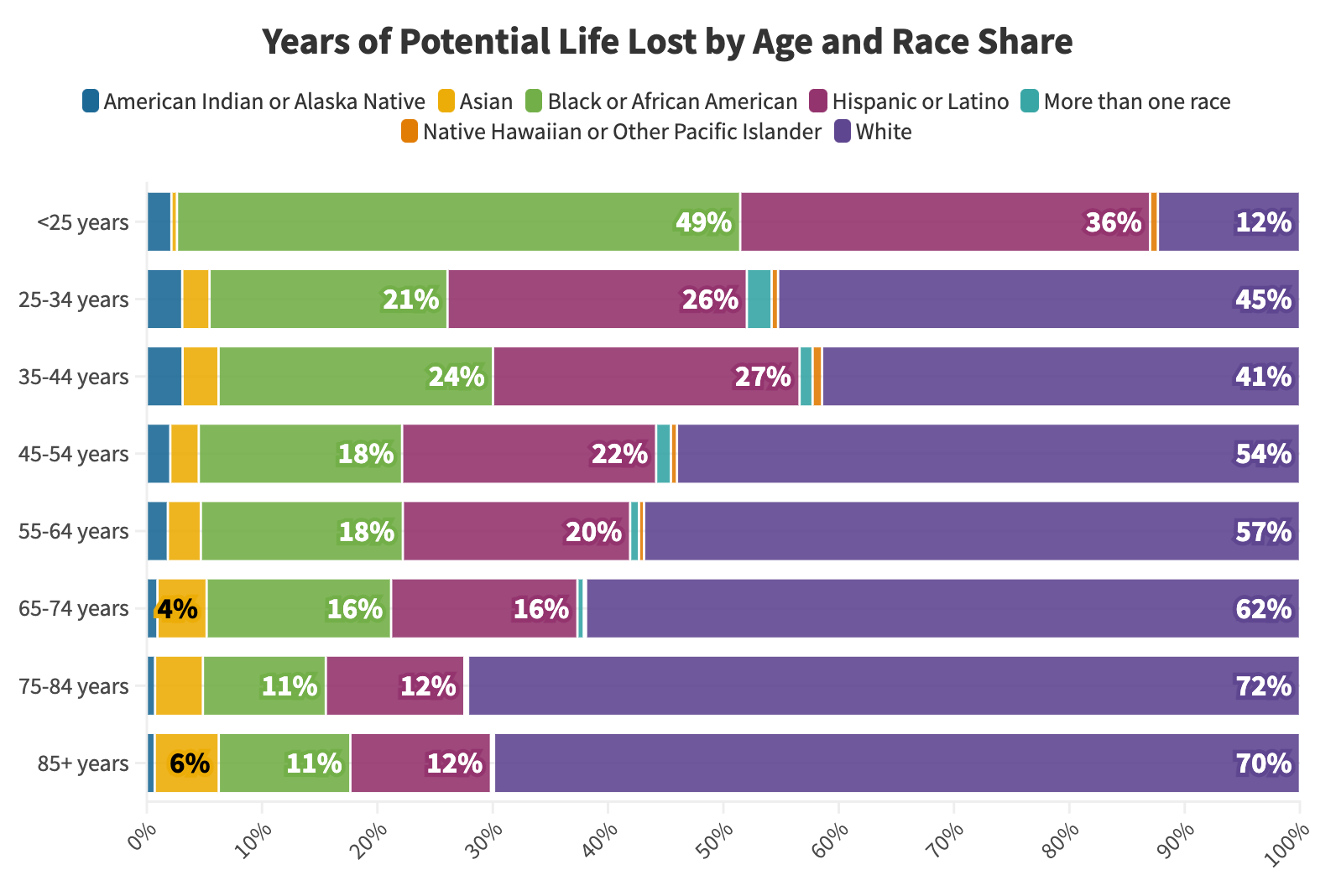

**Figure S5.** Years of potential life lost by race/ethnicity and age group. For NHPI and More than one race categories, YPLL were determined using life expectancy for the AI/AN category, because CDC life tables do not include estimates for these populations.

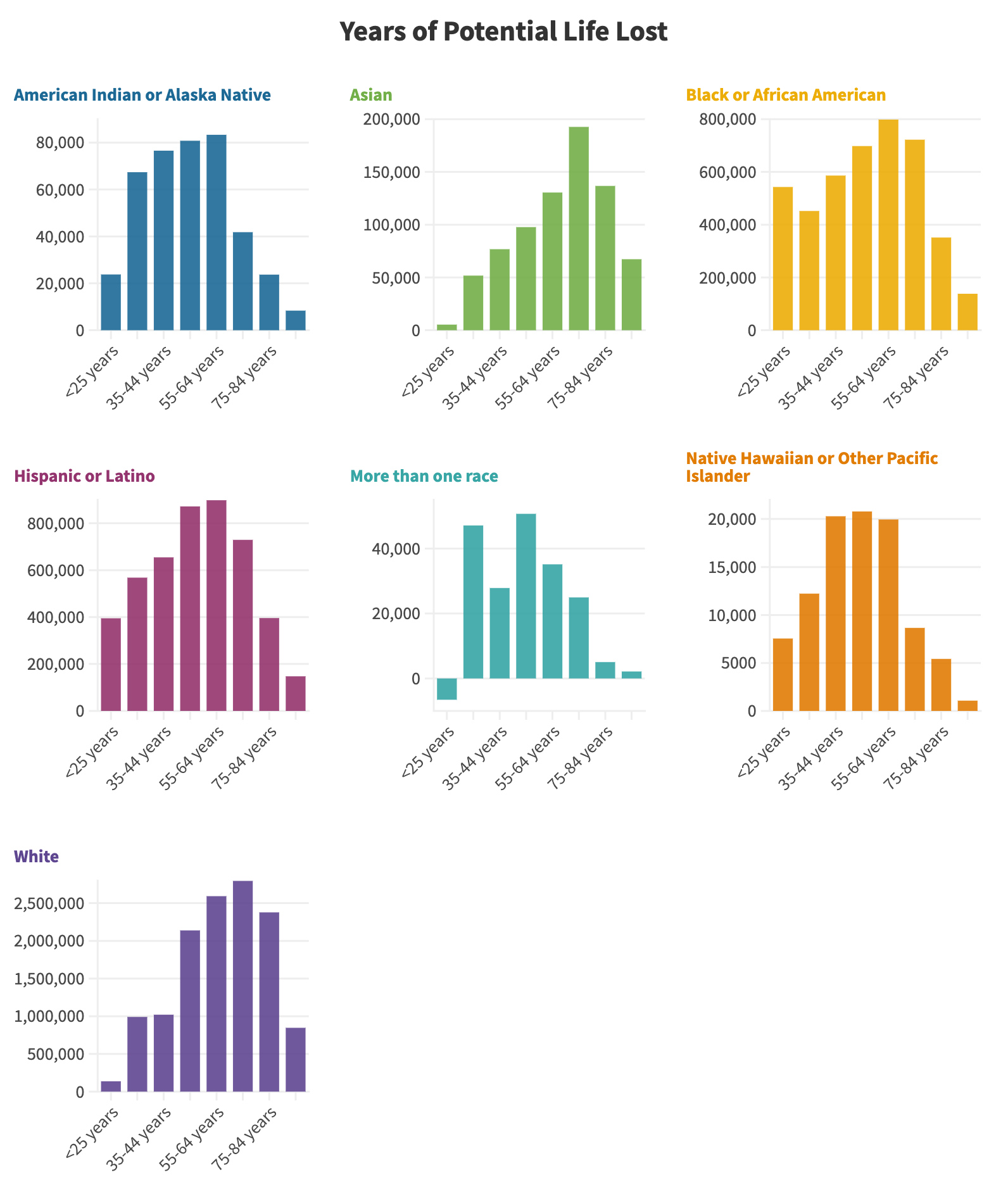

**Figure S6.** Excess mortality per 100,000 persons by race/ethnicity and vaccine period, ages 25-64 years (left panel) and ≥65 years (right panel). 95% confidence intervals are shown in the yellow bars.

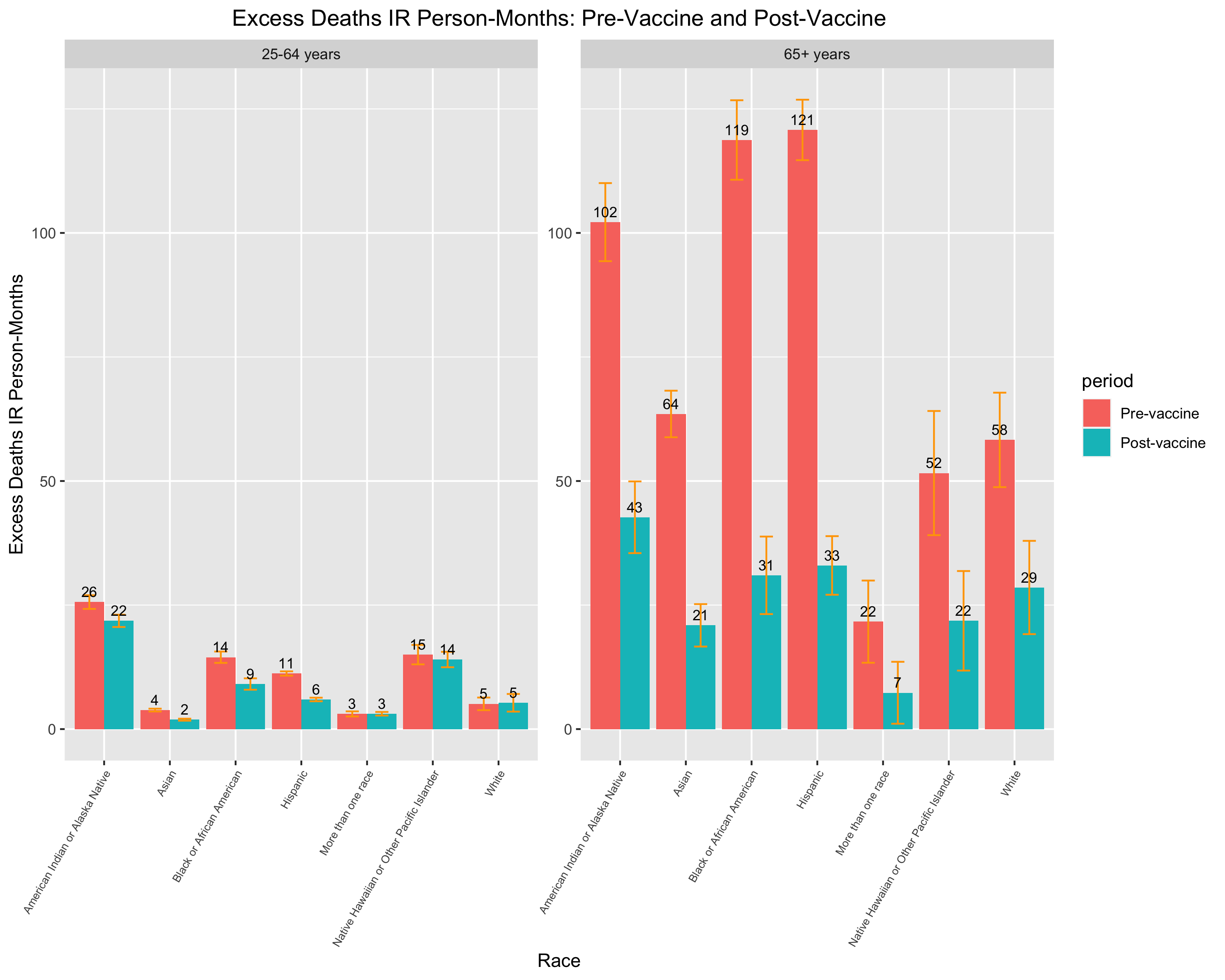

**Figure S7.** Covid-19-specific and all-cause excess mortality per 100,000 persons by race/ethnicity, ages <25 years. Covid-19-specific deaths are not shown in some panels due to suppression or 0 values.

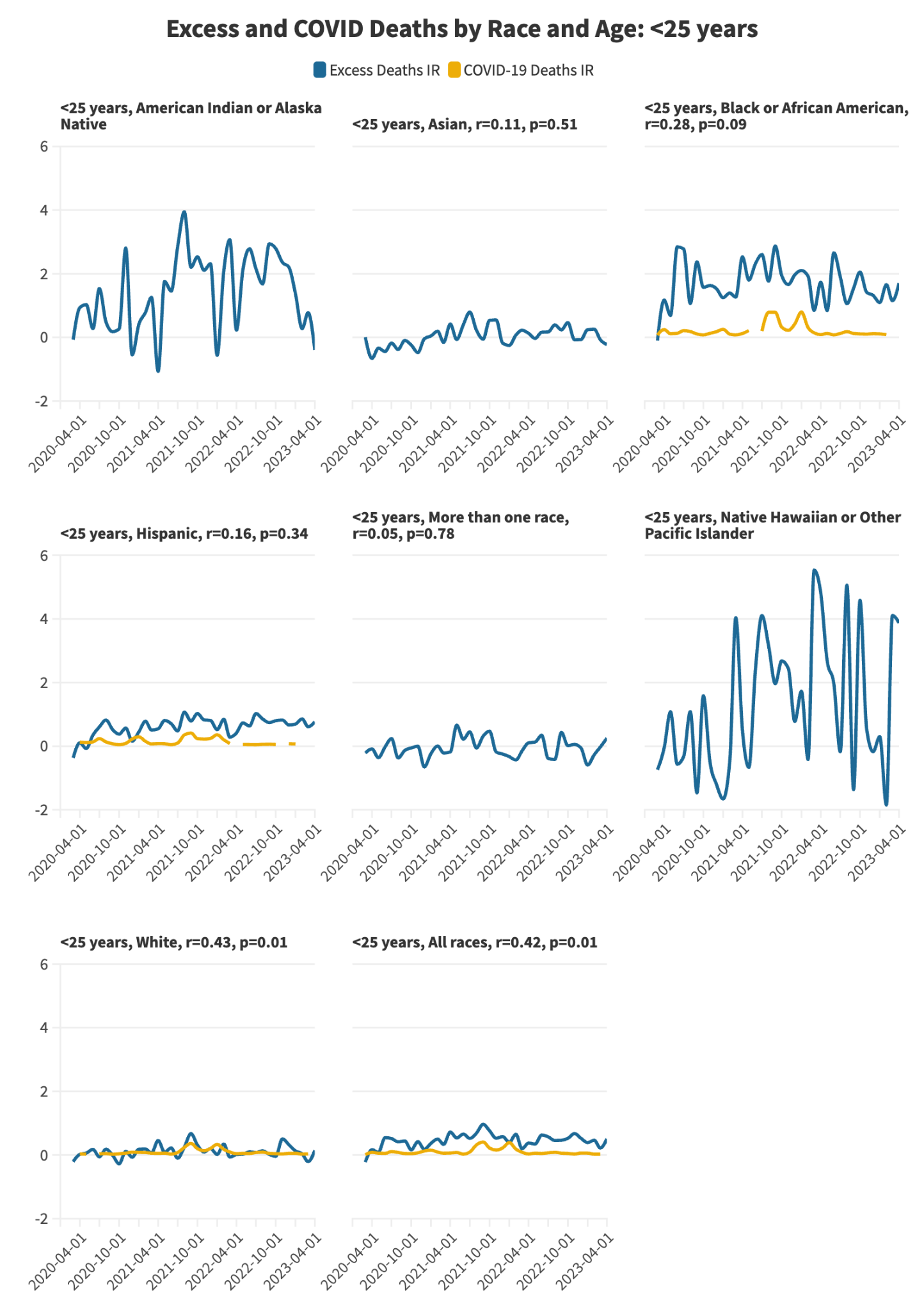

**Figure S8.** Covid-19-specific and all-cause excess mortality per 100,000 persons by race/ethnicity, ages 25-64 years.

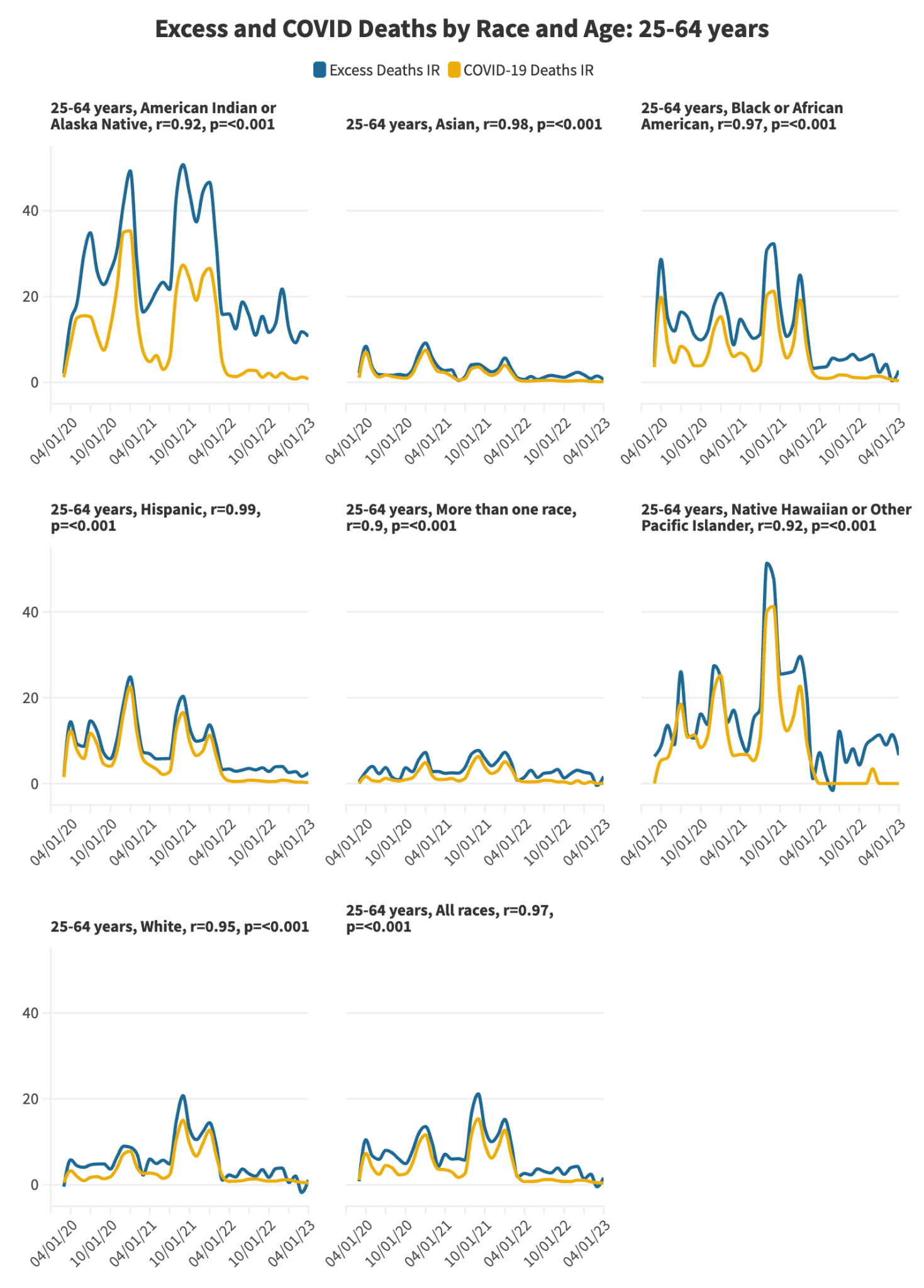

**Figure S9.** Covid-19-specific and all-cause excess mortality per 100,000 persons by race/ethnicity, ages ≥65 years.

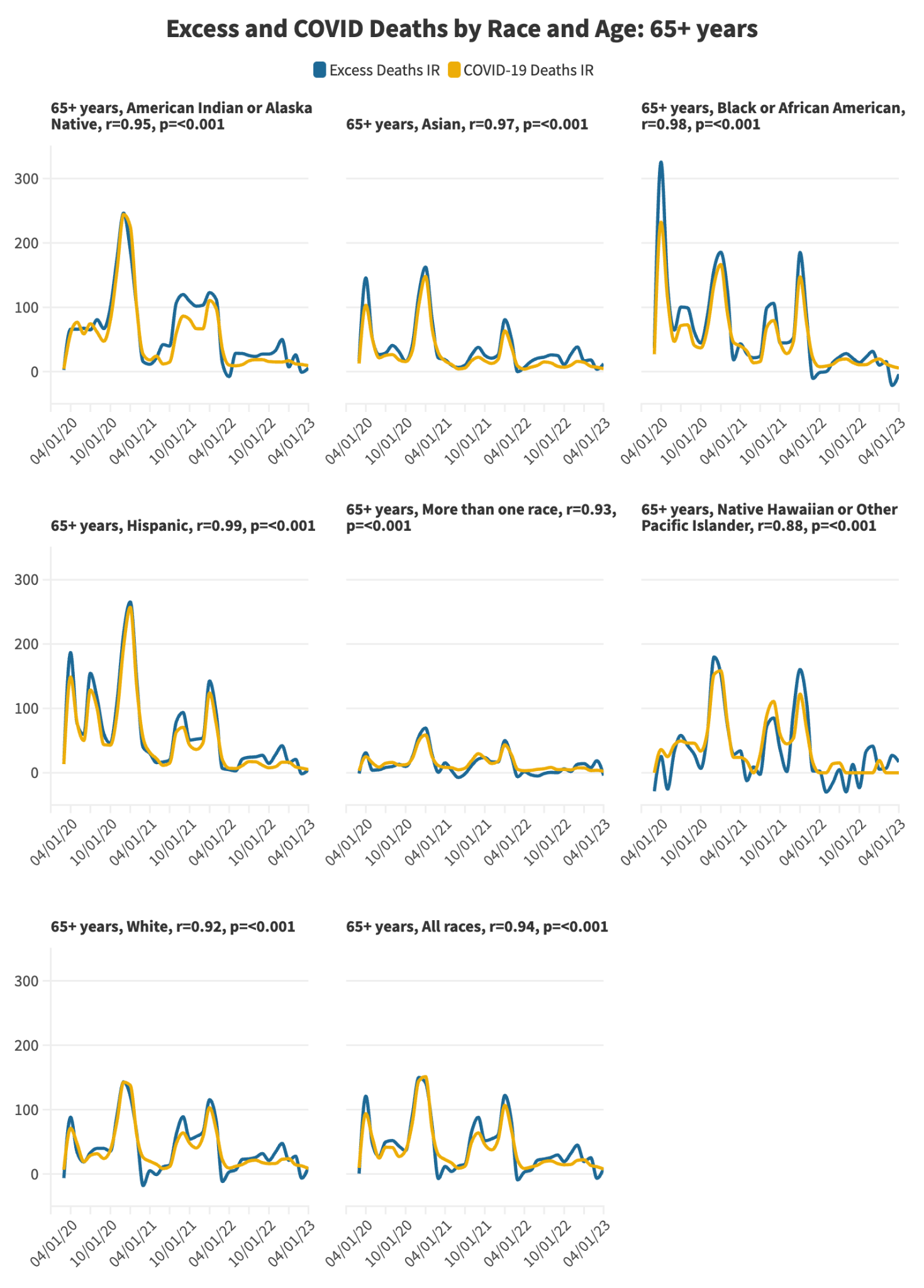

**Table S6.** Pearson correlation between all-cause excess mortality and Covid-19-specific mortality (multiple cause of death) by race/ethnicity and age group, with corresponding p values.

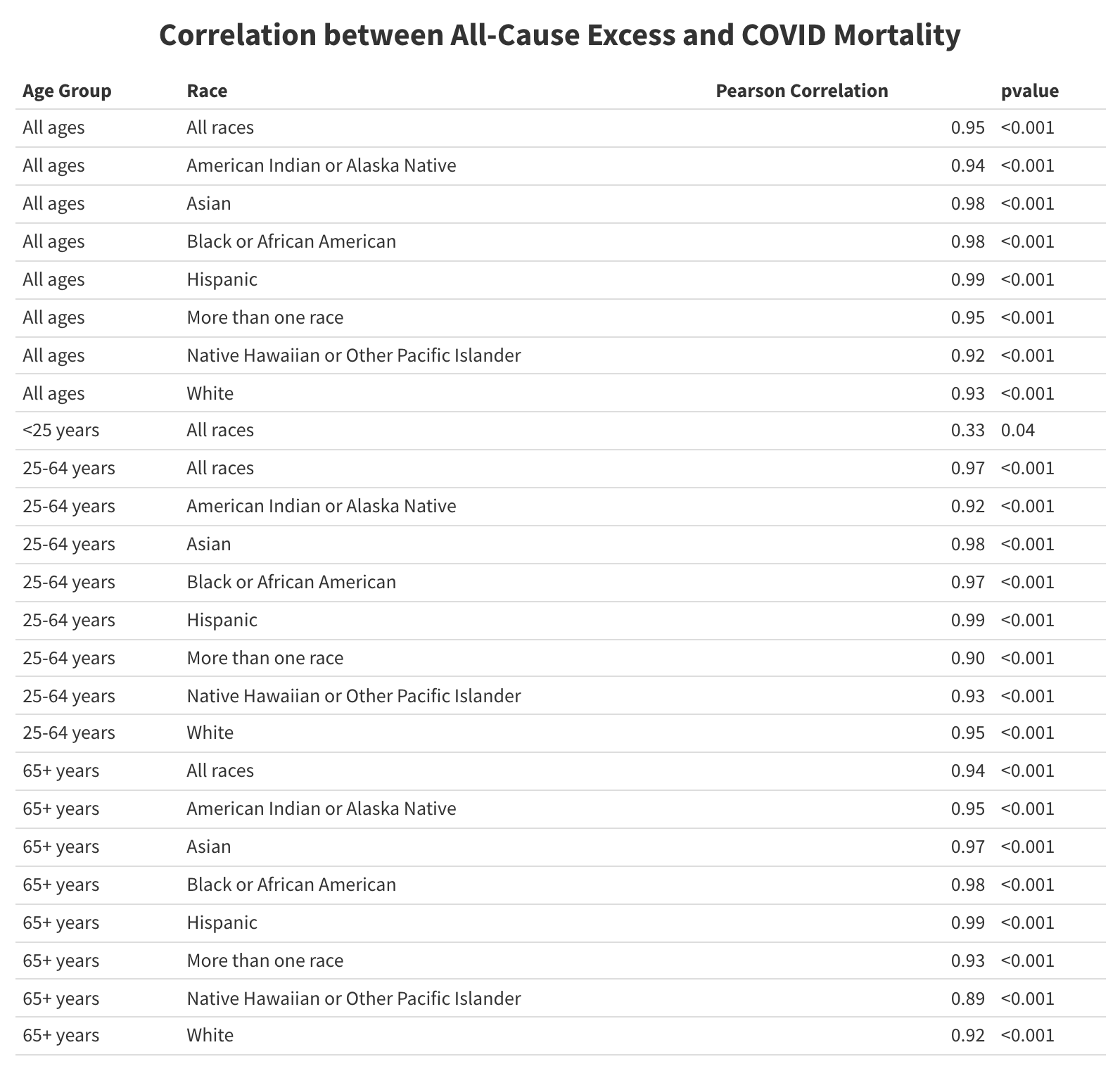

**Figure S10. Changes in cause-specific mortality and correlation to Covid-19-specific mortality.** Observed deaths per 100,000 persons by race/ethnicity (all ages) and UCD – ICD Chapter. The pandemic period cause-specific incident rate per 100,000 persons is shown in the left column; the middle and right columns show the correlation between the row cause and Covid-19-specific deaths and the corresponding p value, respectively. *Analysis*: In the all-ages category, diseases of the circulatory system and endocrine, nutritional, and metabolic diseases (including diabetes) were most frequently correlated with Covid-19 mortality (see also, Figure 3) and within the age-specific groups (Table S8). In the all-ages analysis, strong correlations were observed between cause-specific mortality and Covid-19 mortality for endocrine, nutritional, and metabolic diseases (Black) and diseases of the genitourinary system (White). Strong correlations with Covid-19 mortality were also seen in the Black and White populations for mental and behavioral disorders, diseases of the nervous system (including Alzheimer’s), and circulatory system diseases.

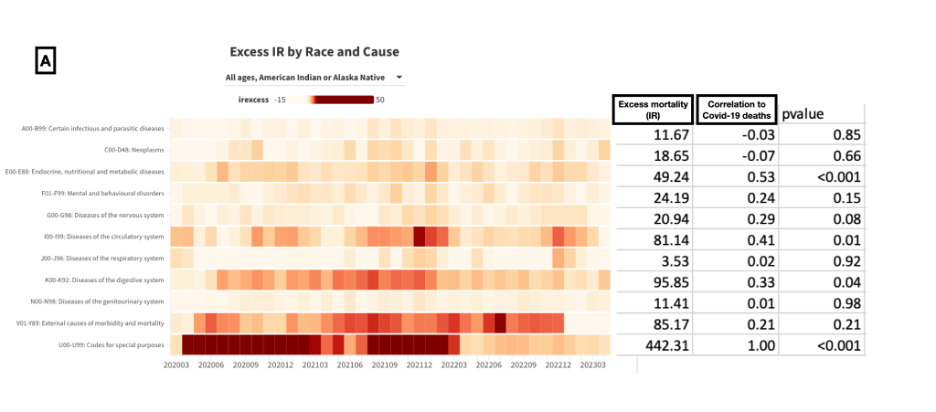

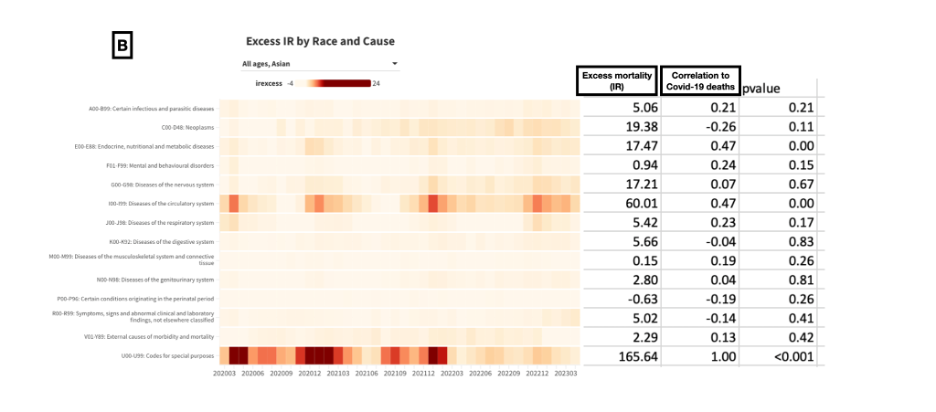

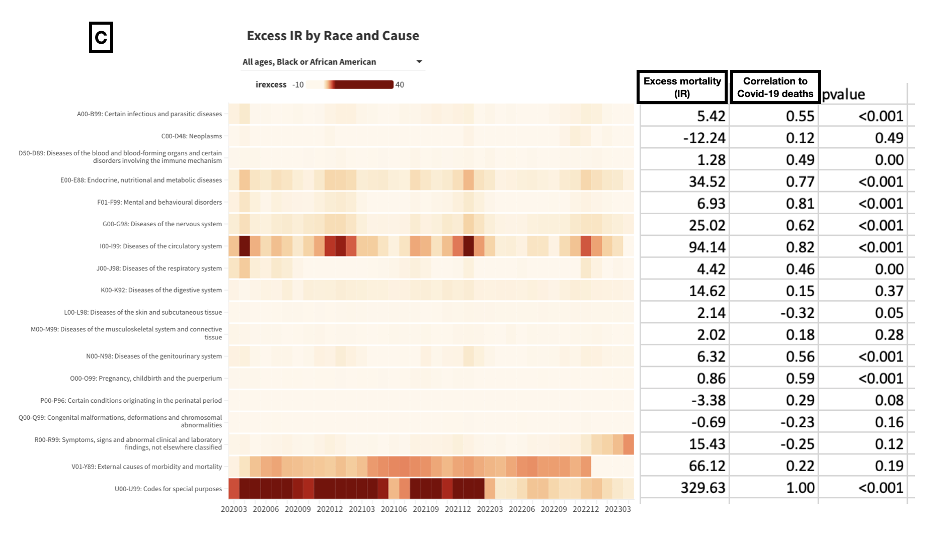

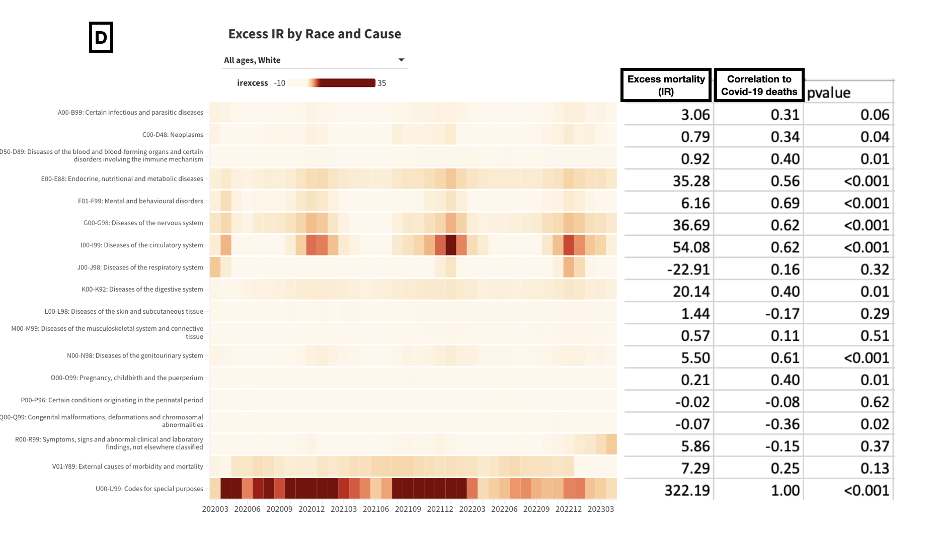

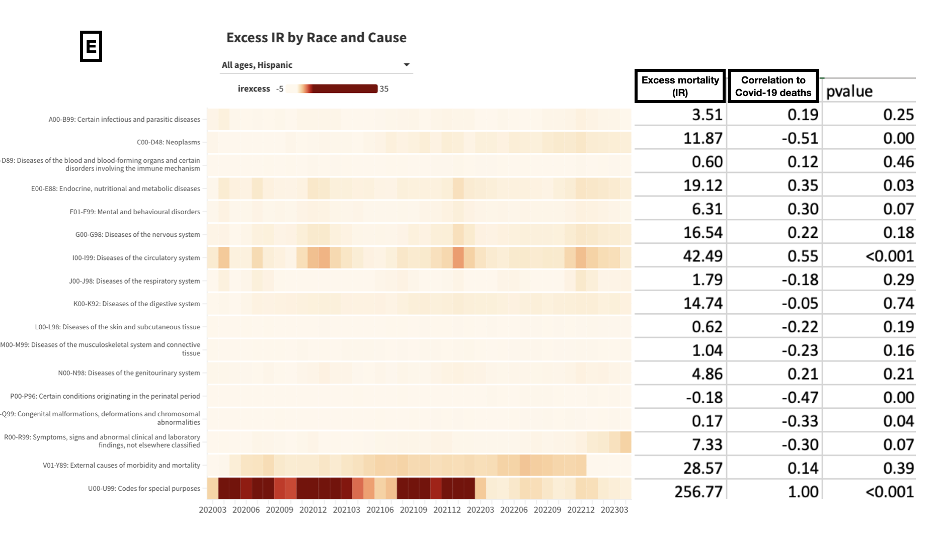

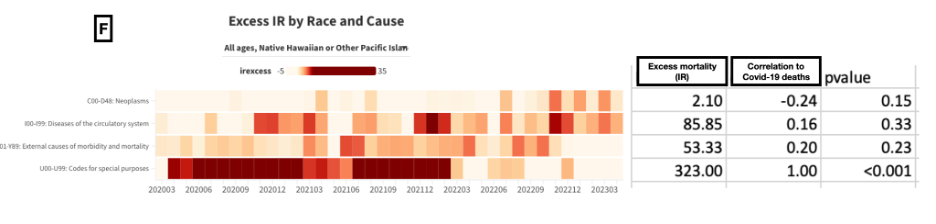

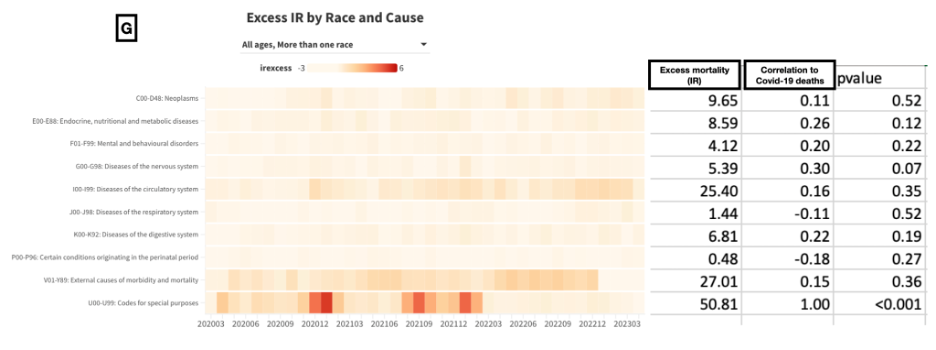

**Figure S11.** Observed deaths per 100,000 persons by race/ethnicity (ages <25 years) and UCD – ICD Chapter. The pandemic period cause-specific incident rate per 100,000 persons is shown in the left column; the middle and right columns show the correlation between the row cause and (if possible) Covid-19-specific deaths and the corresponding p value, respectively.

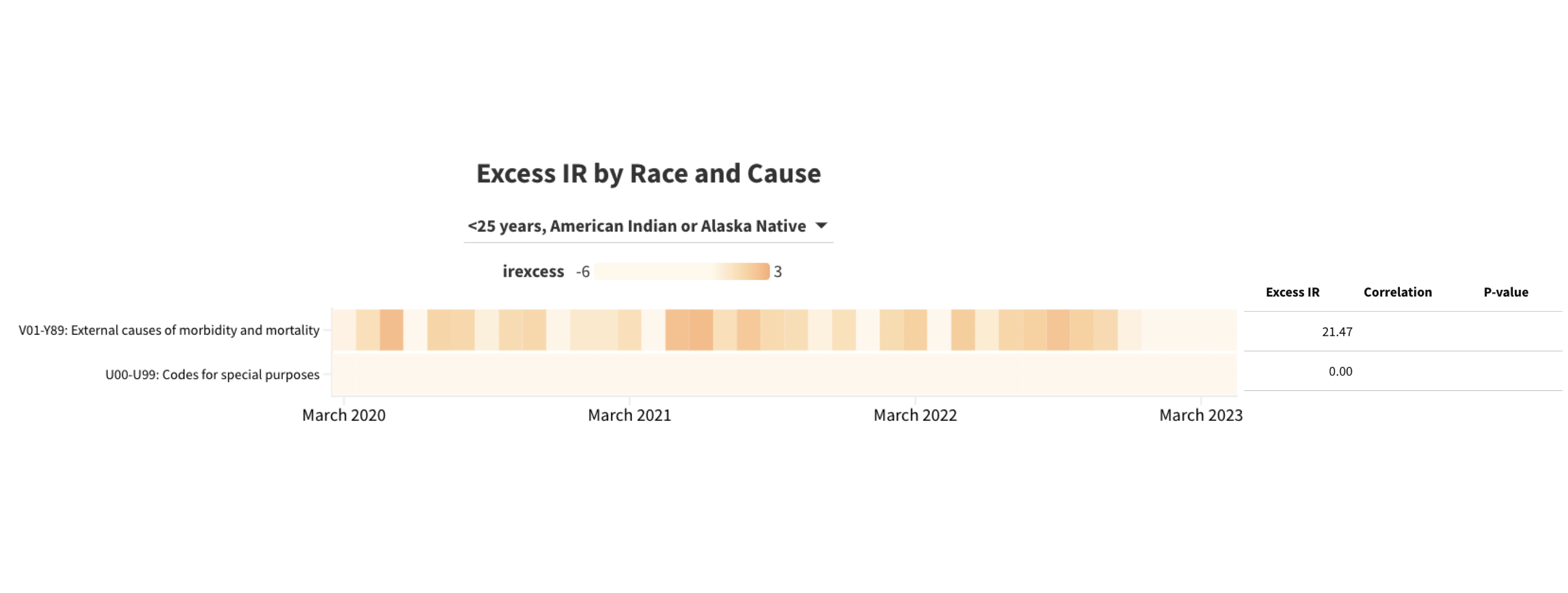

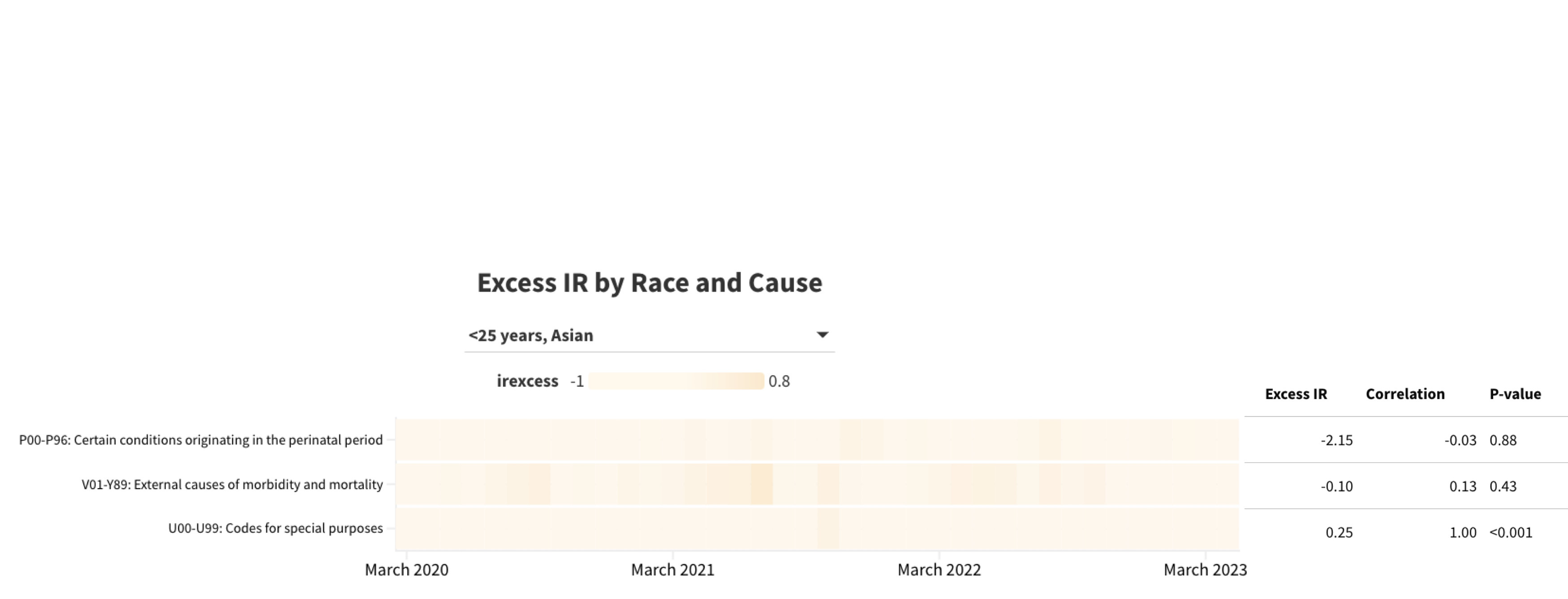

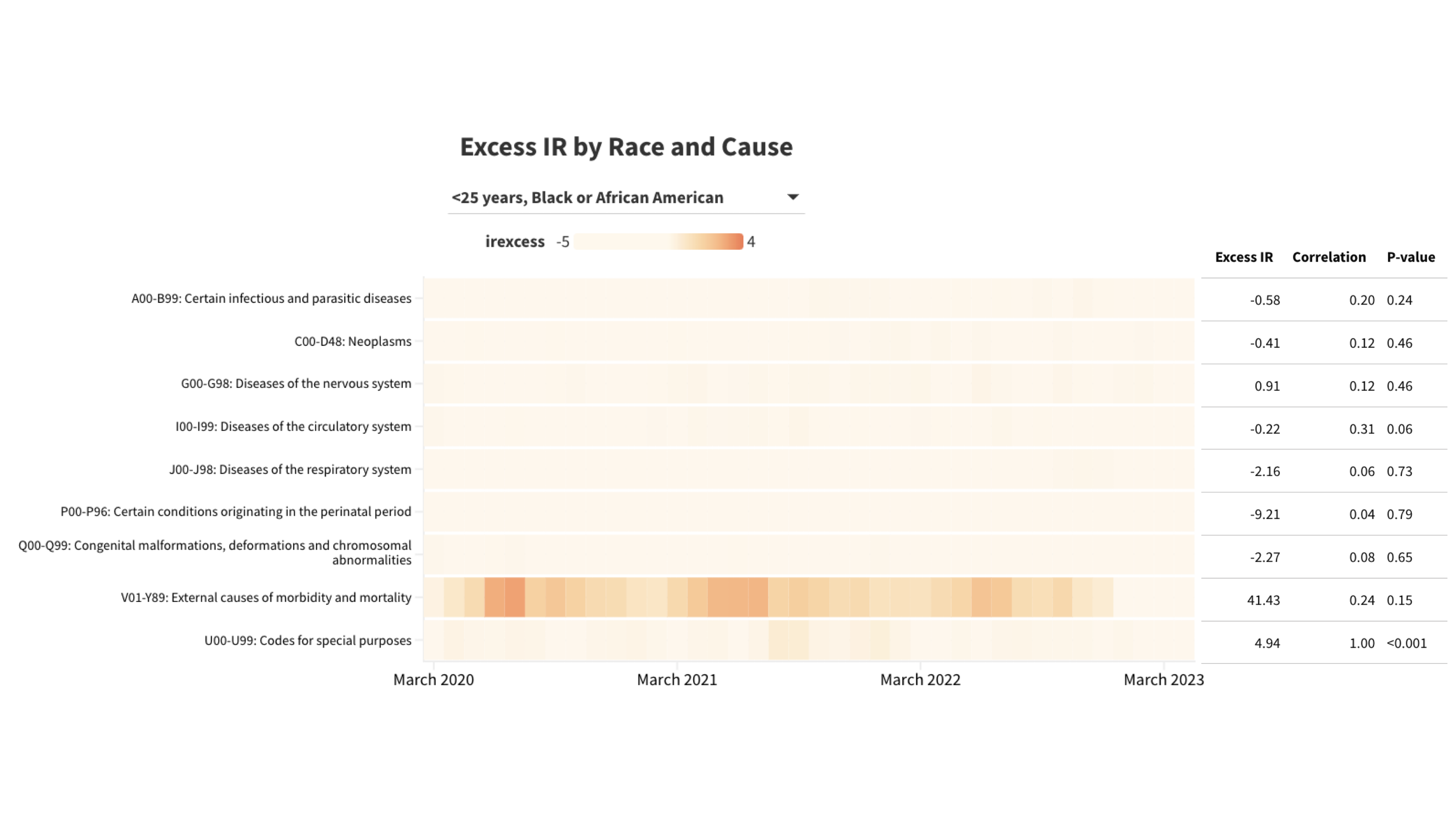

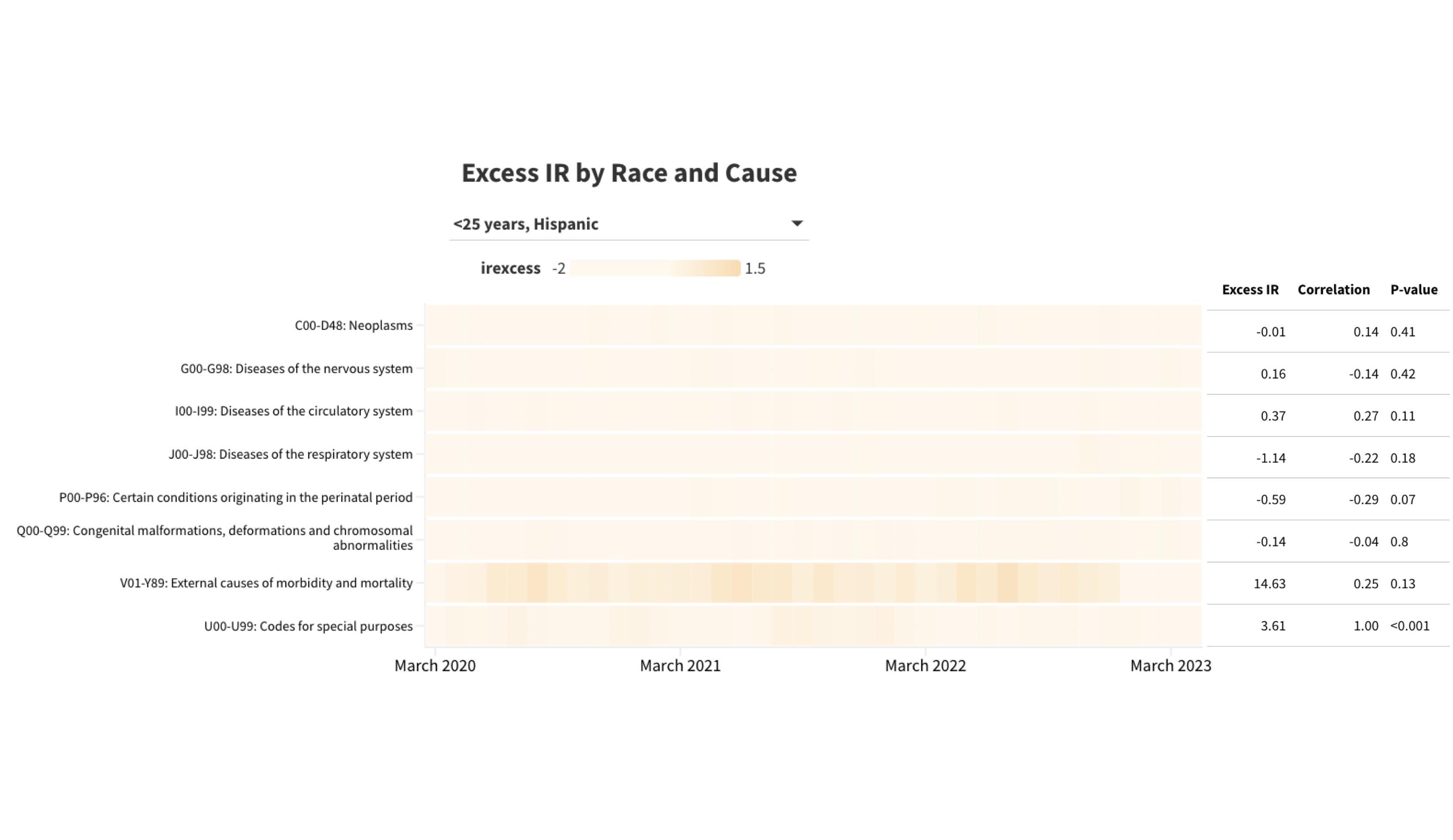

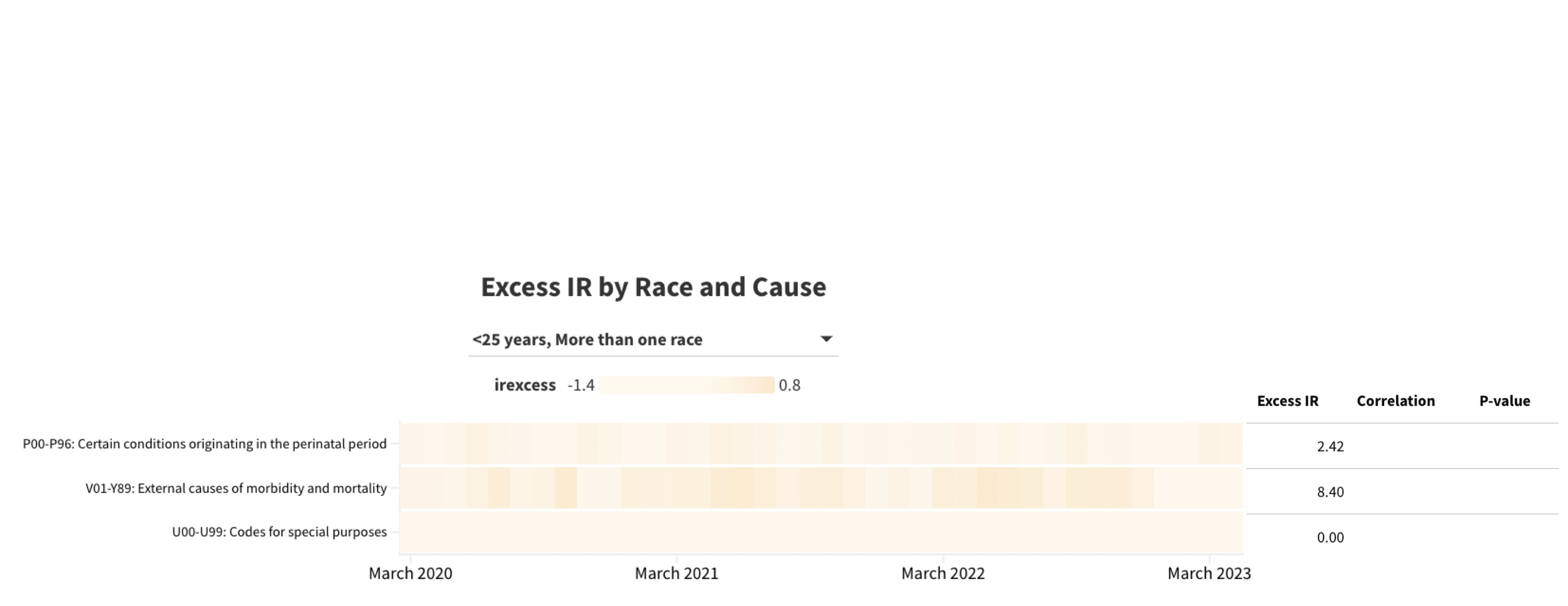

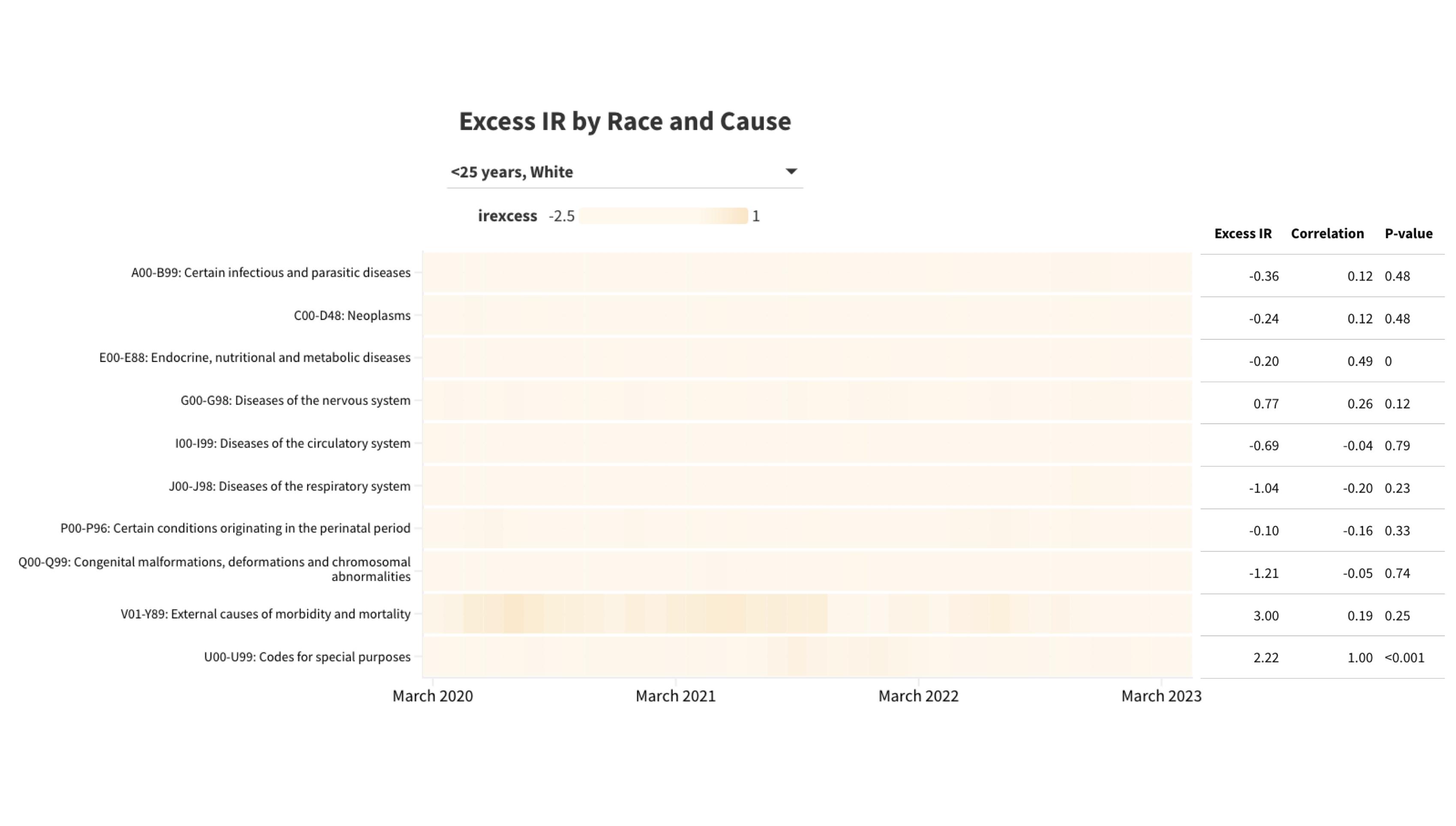

**Figure S12.** Observed deaths per 100,000 persons by race/ethnicity (ages 25-64) and UCD – ICD Chapter. The pandemic period cause-specific incident rate per 100,000 persons is shown in the left column; the middle and right columns show the correlation between the row cause and Covid-19-specific deaths and the corresponding p value, respectively.
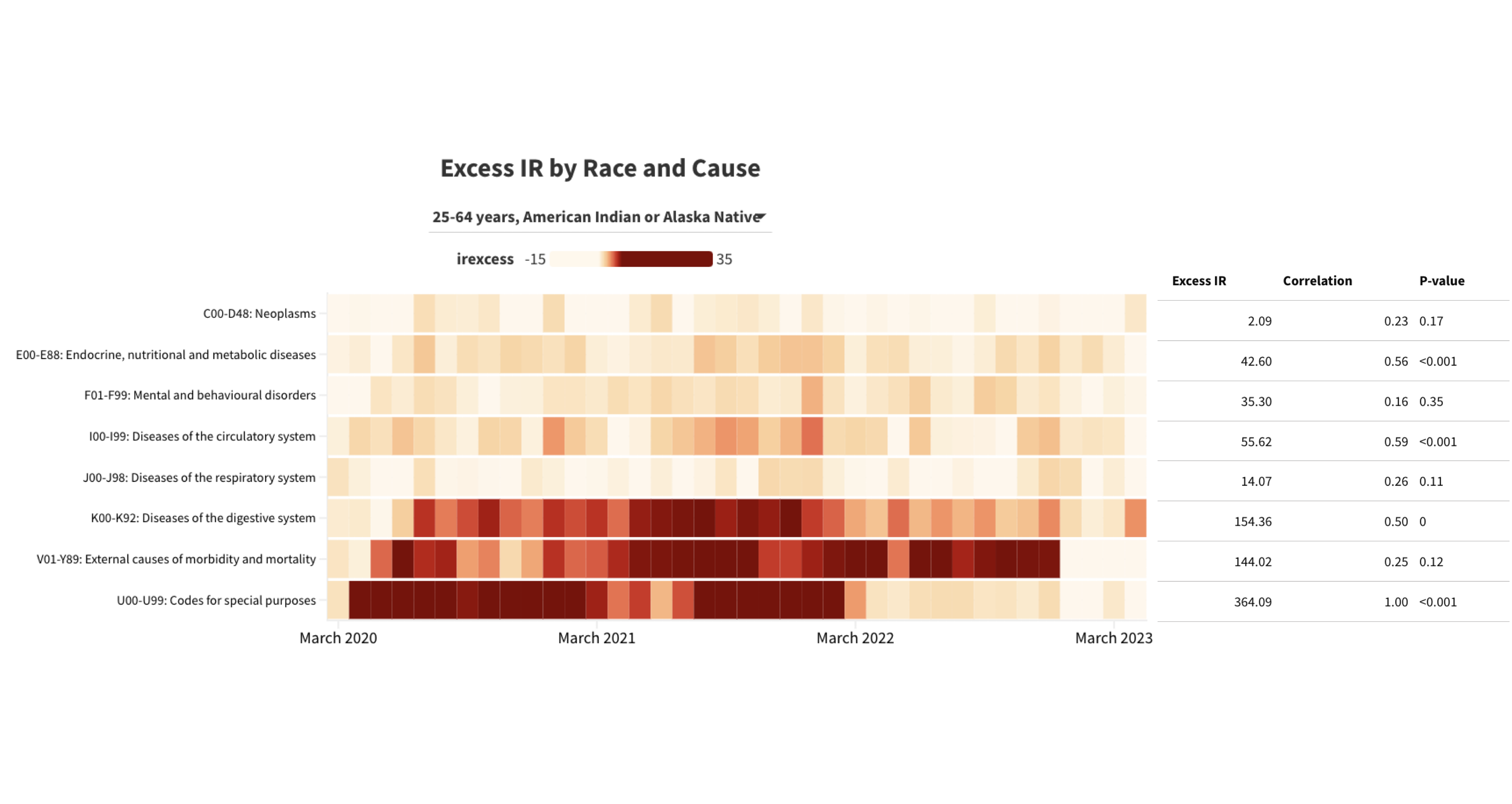

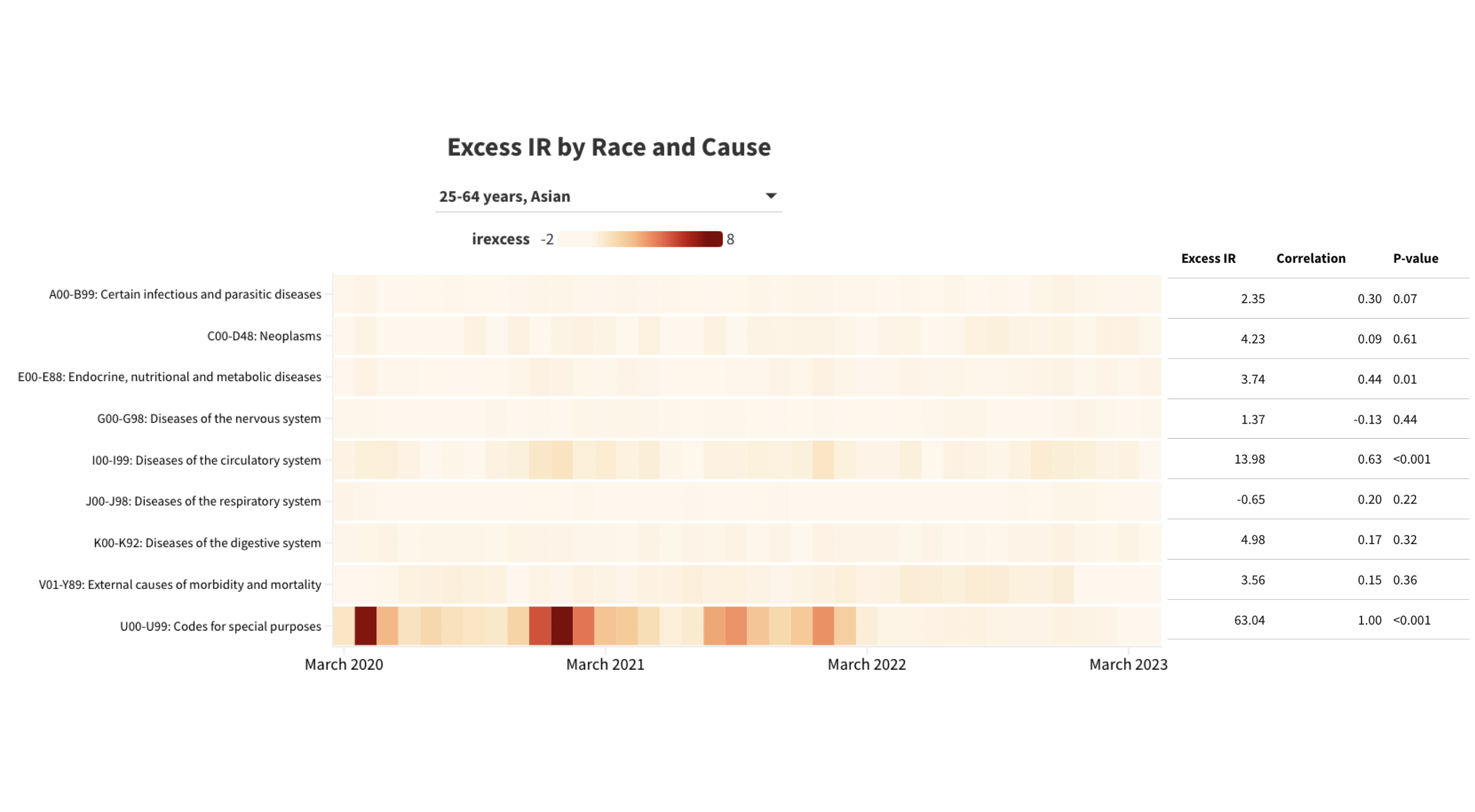

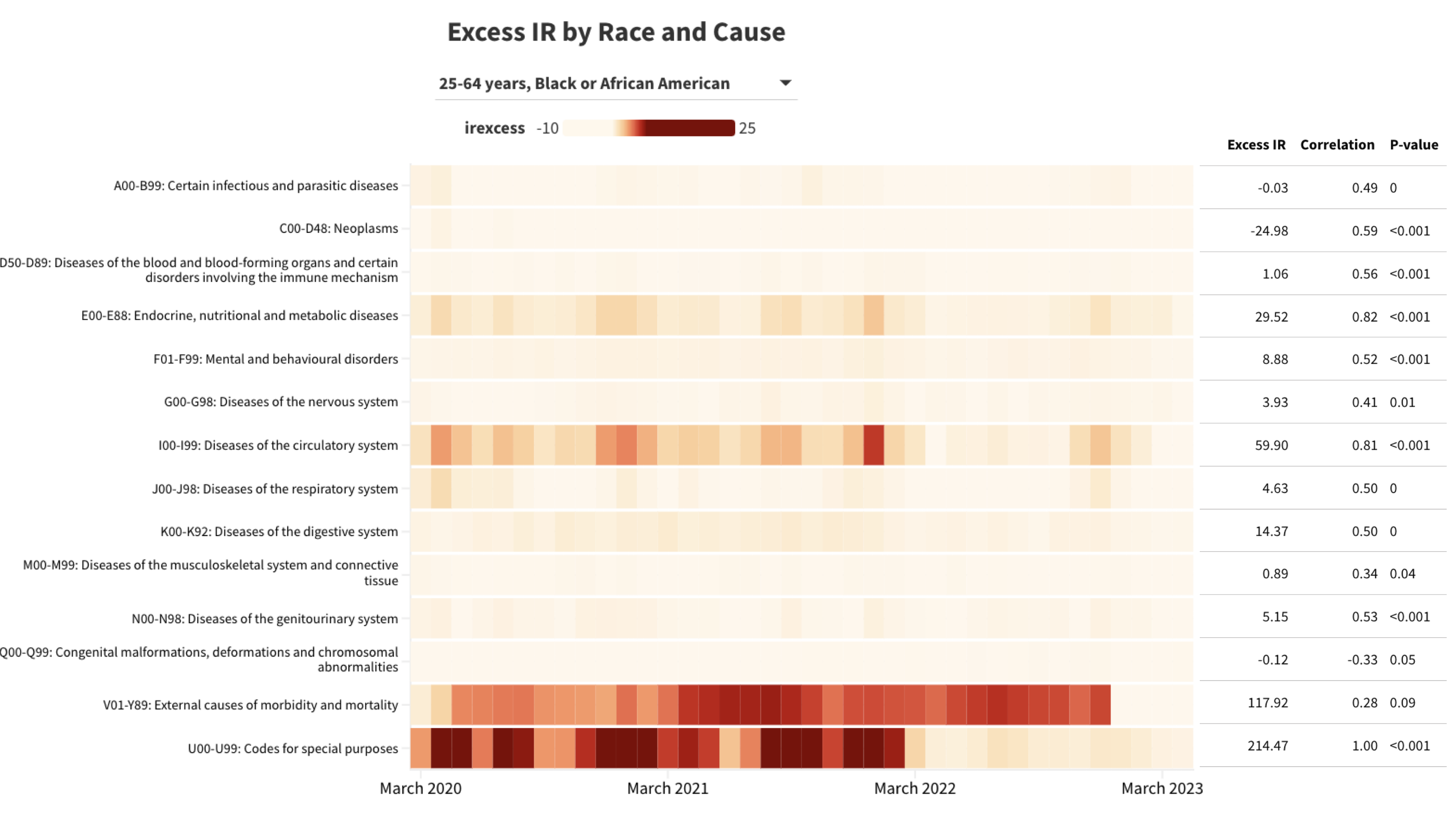

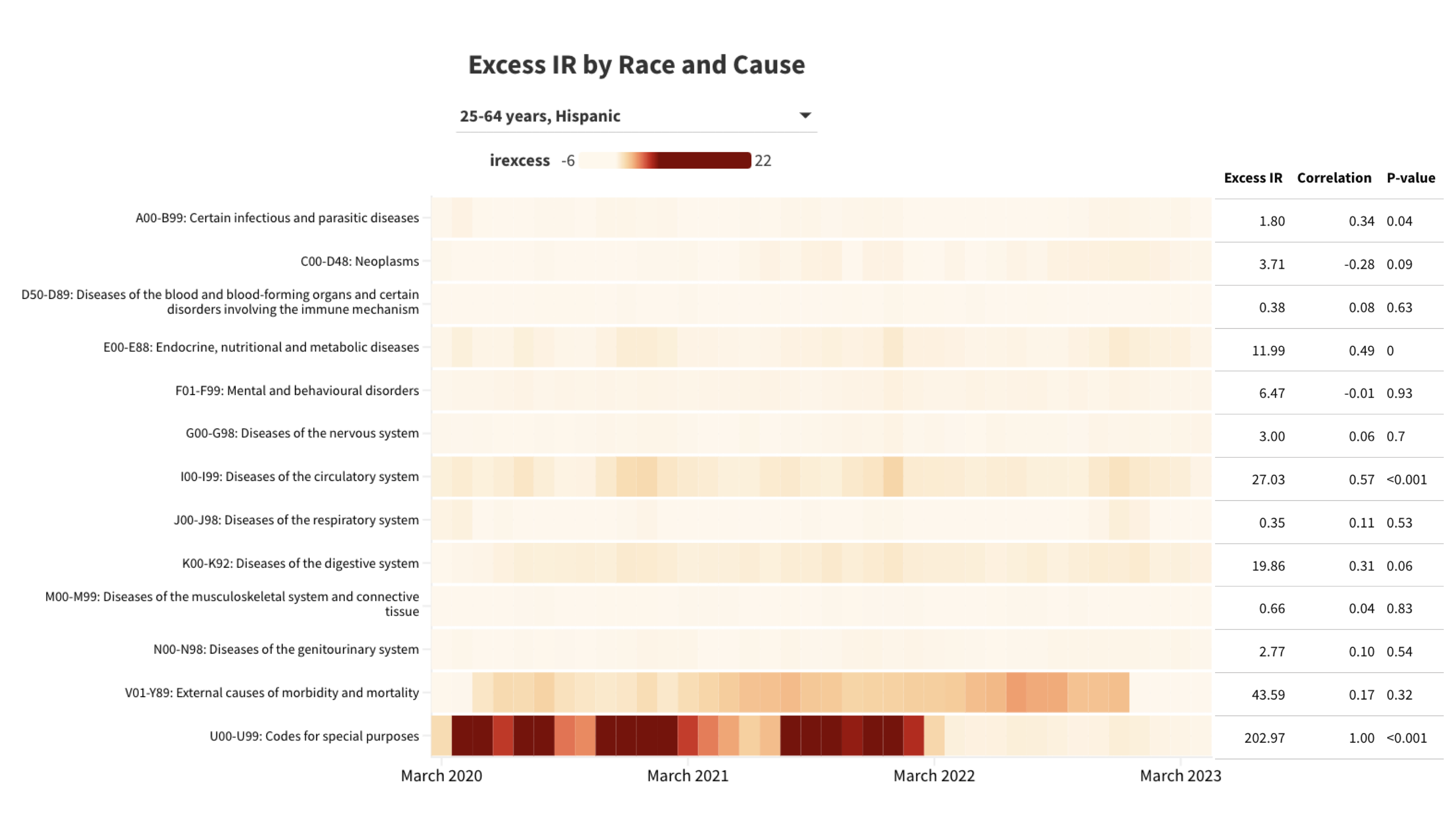

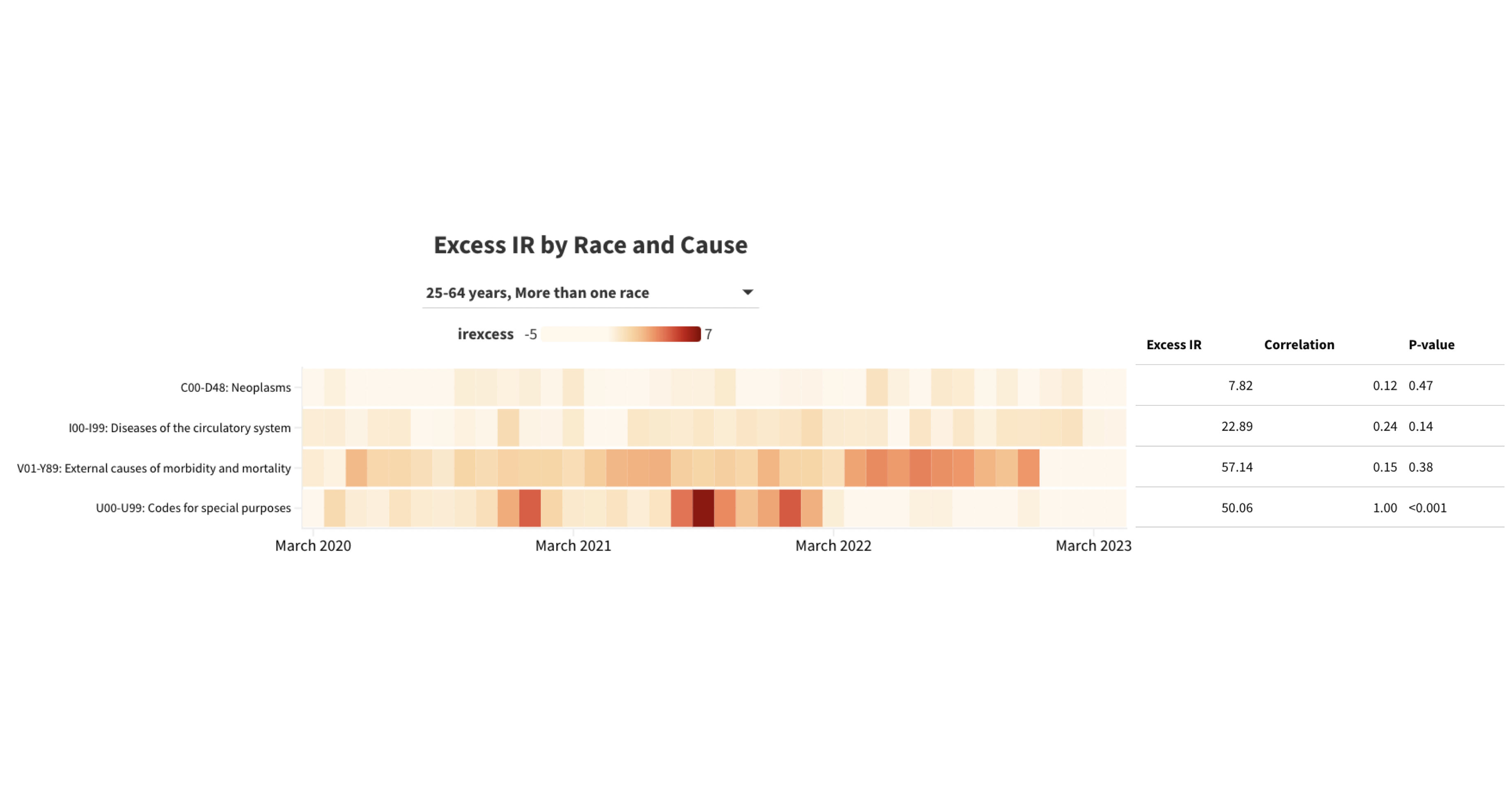

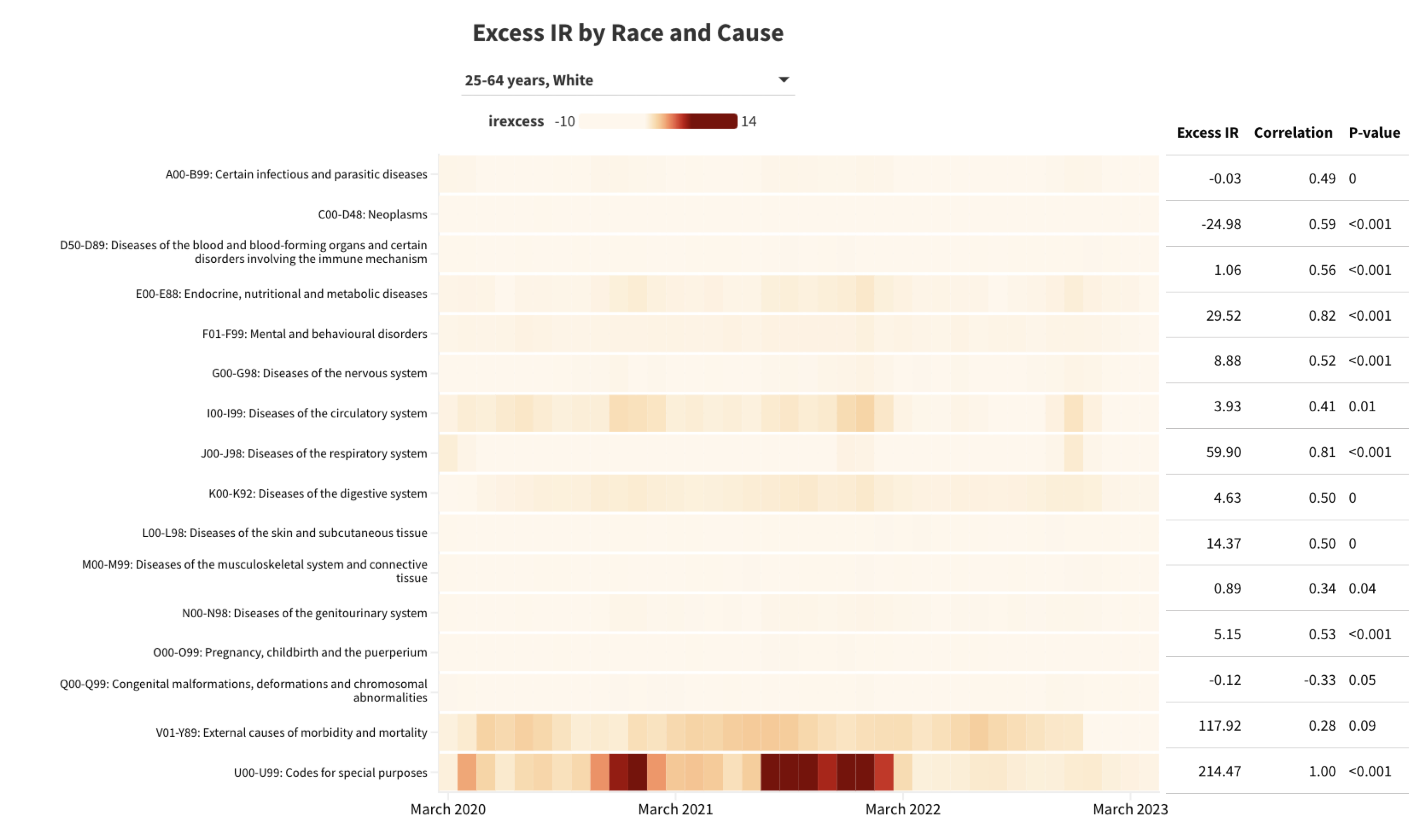

**Figure S13.** Observed deaths per 100,000 persons by race/ethnicity (ages ≥65 years) and UCD – ICD Chapter. The pandemic period cause-specific incident rate per 100,000 persons is shown in the left column; the middle and right columns show the correlation between the row cause and Covid-19-specific deaths and the corresponding p value, respectively.

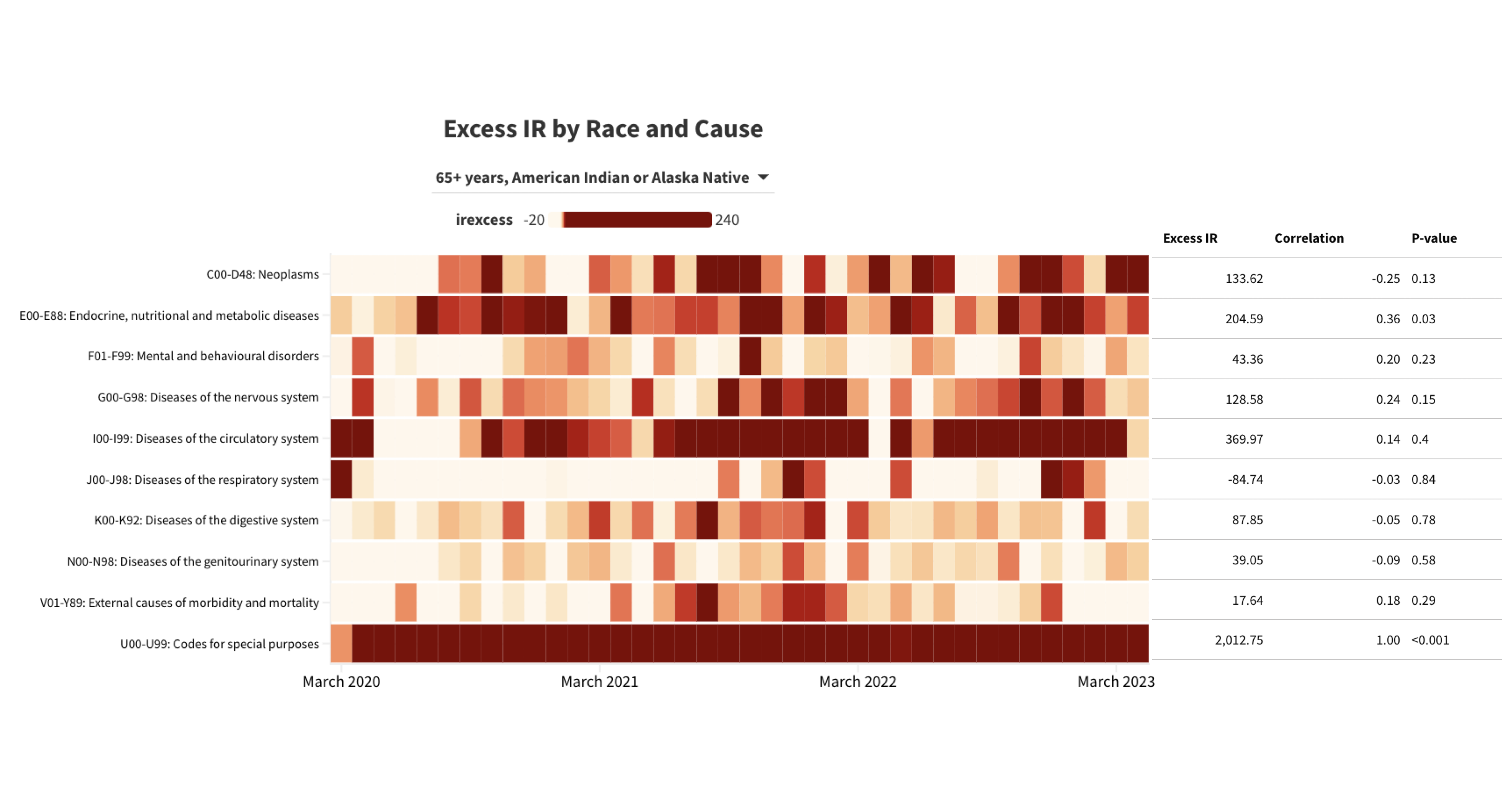

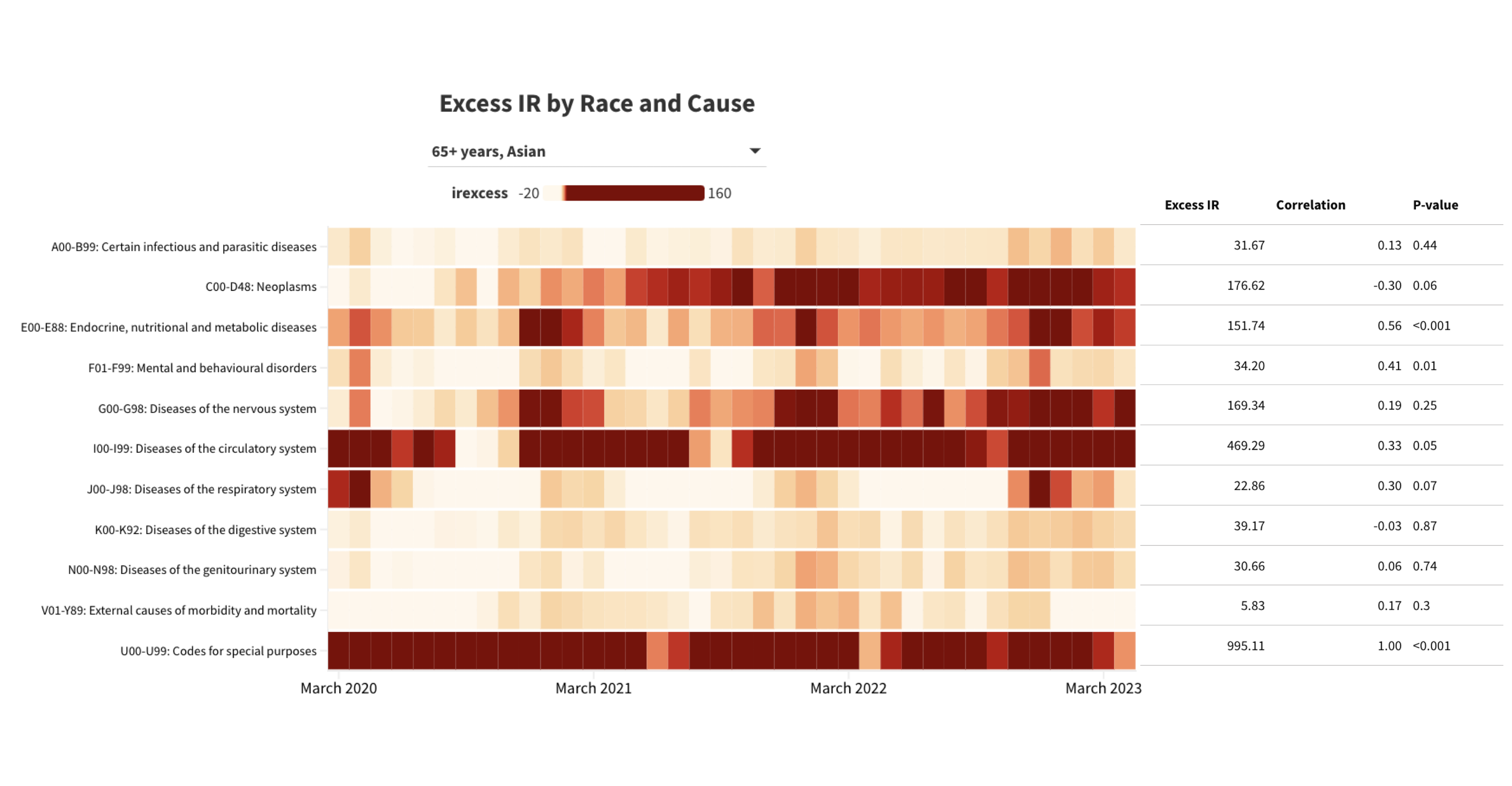

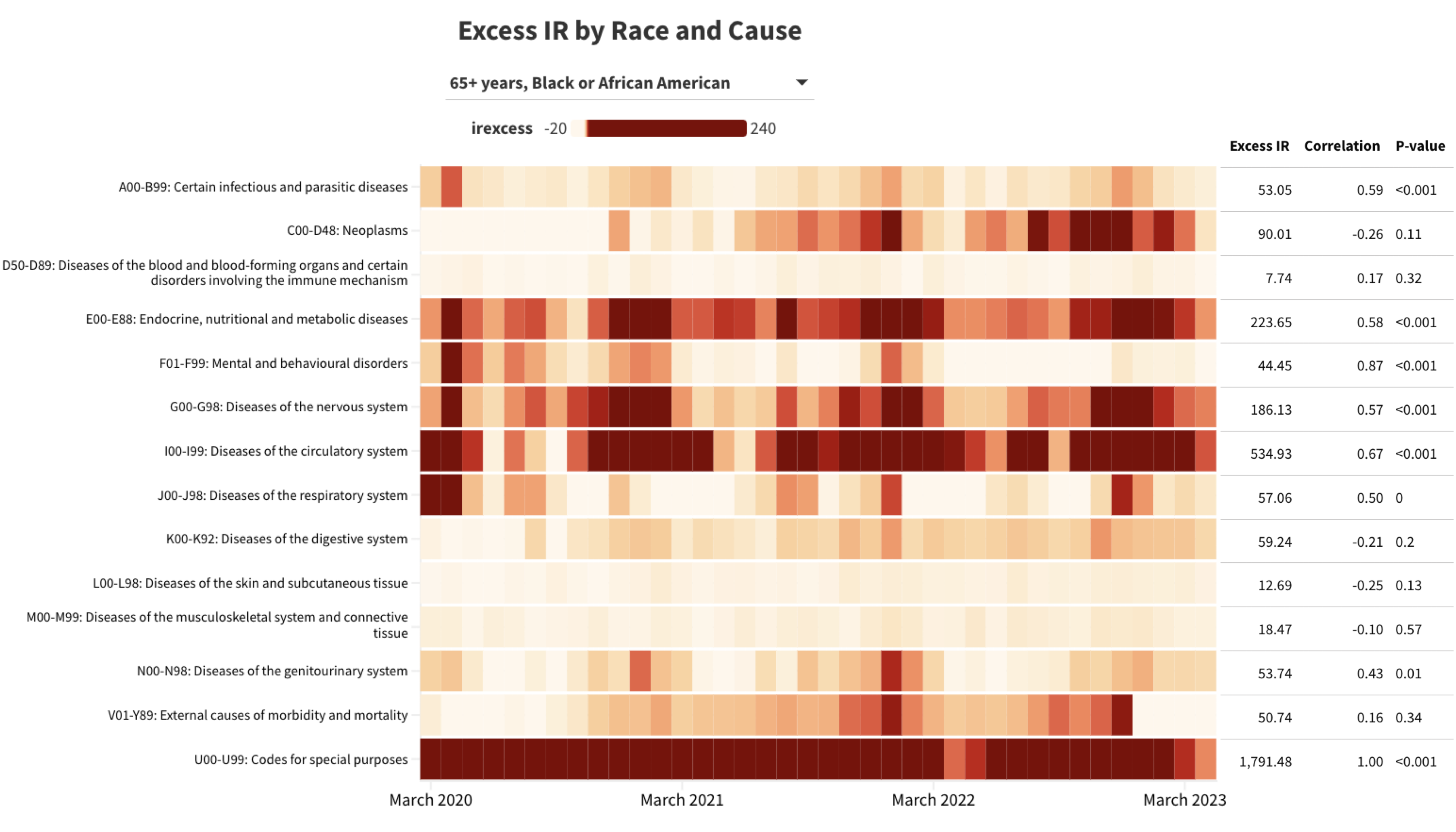

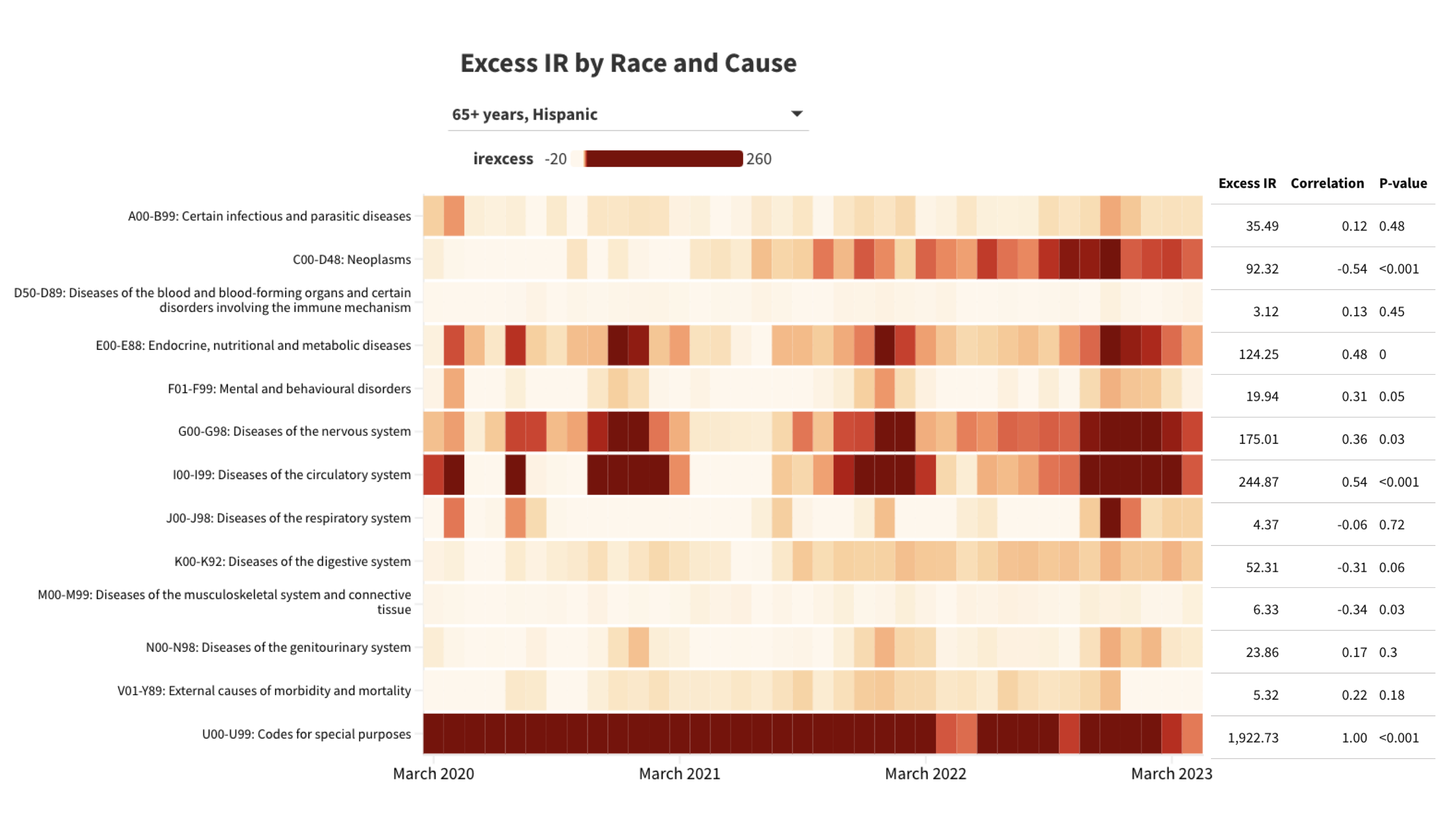

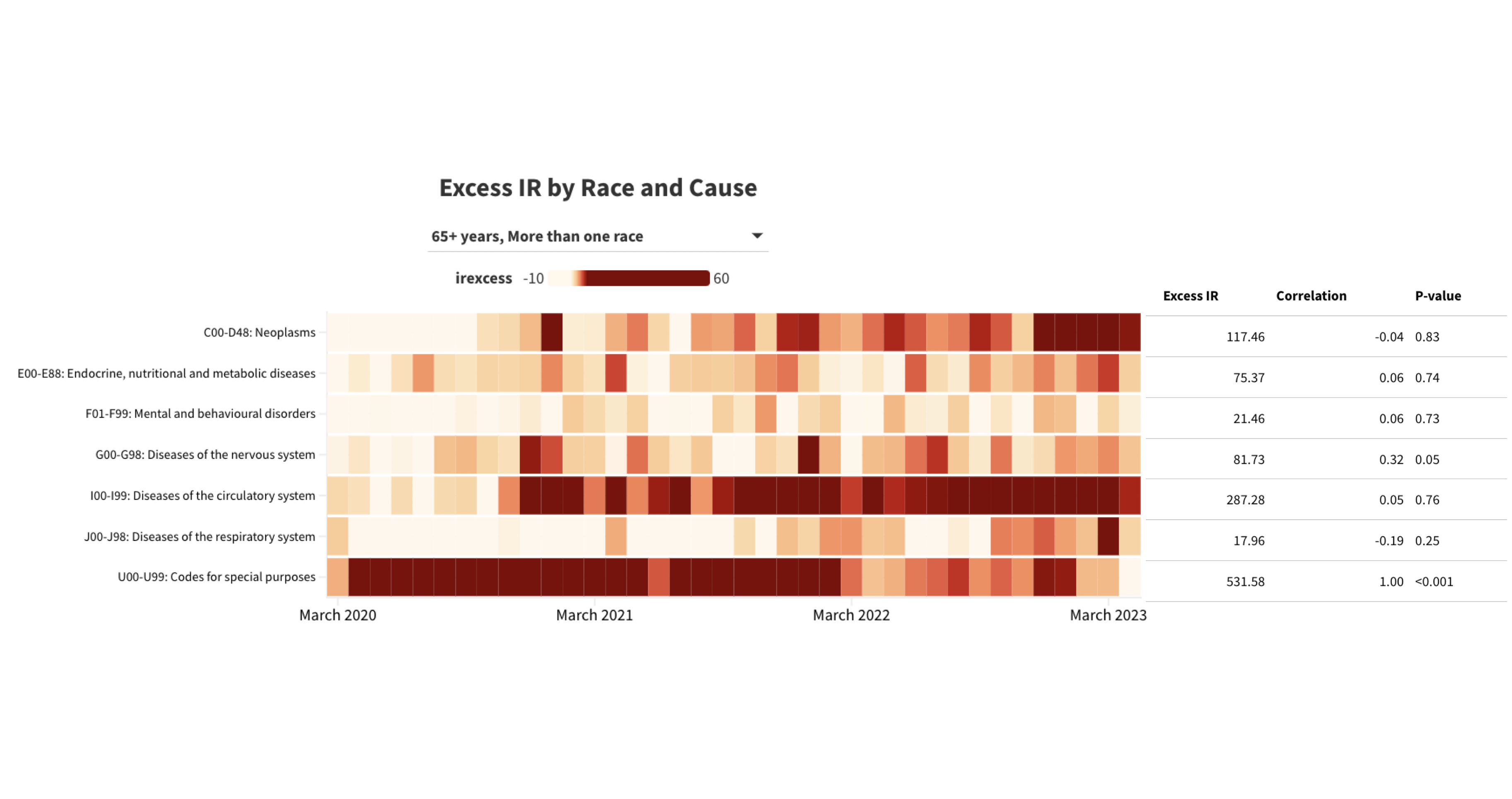

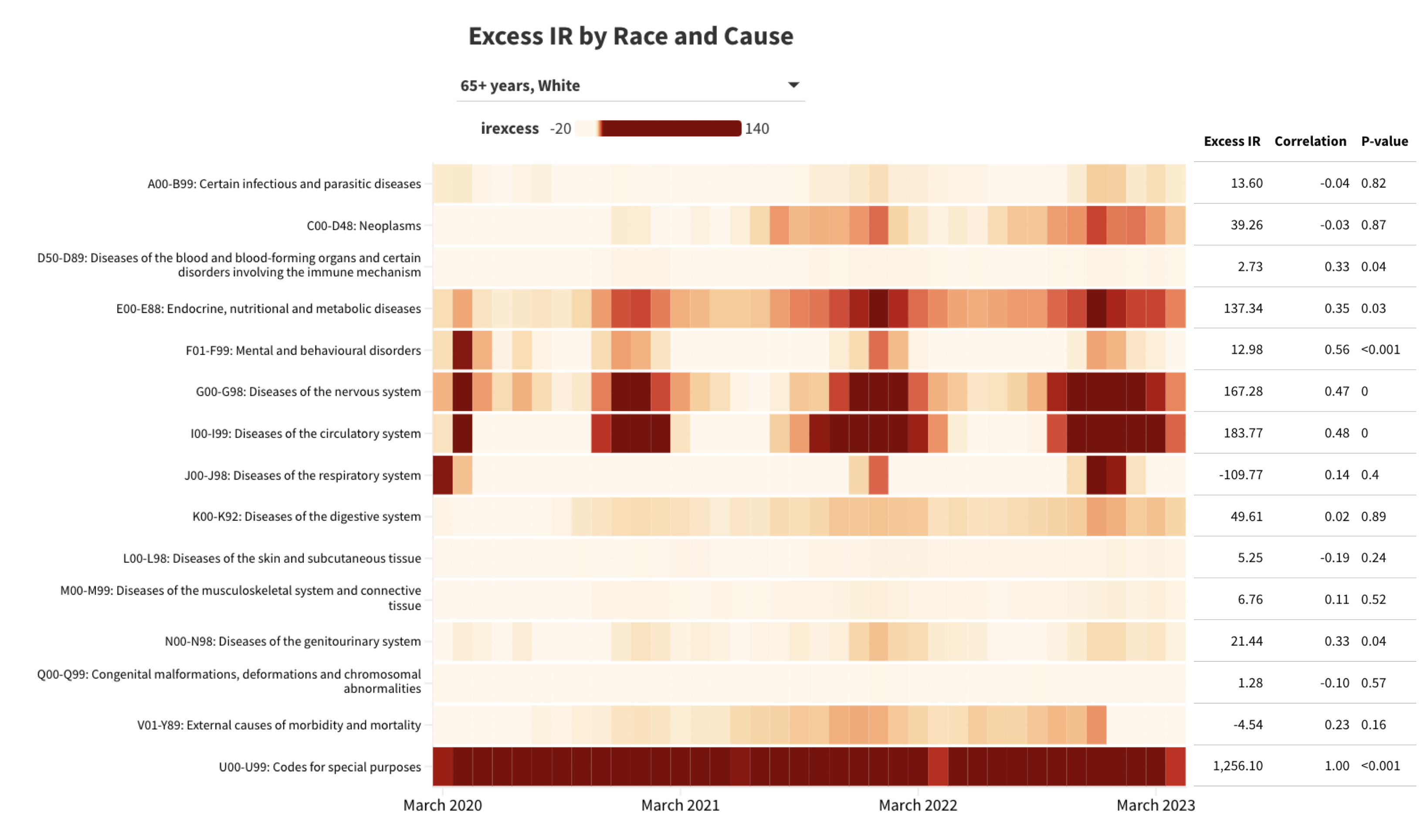

**Table S7**

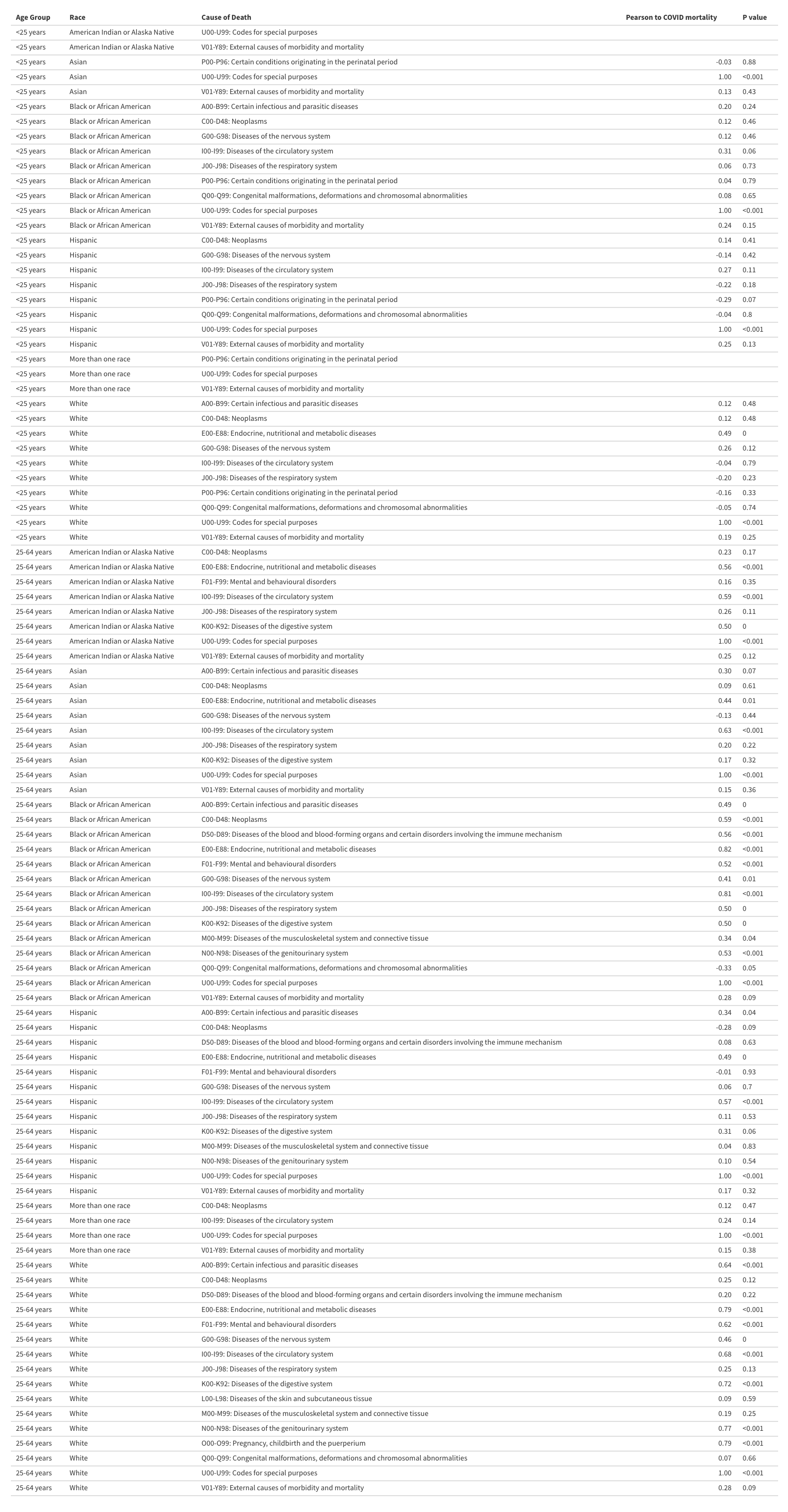

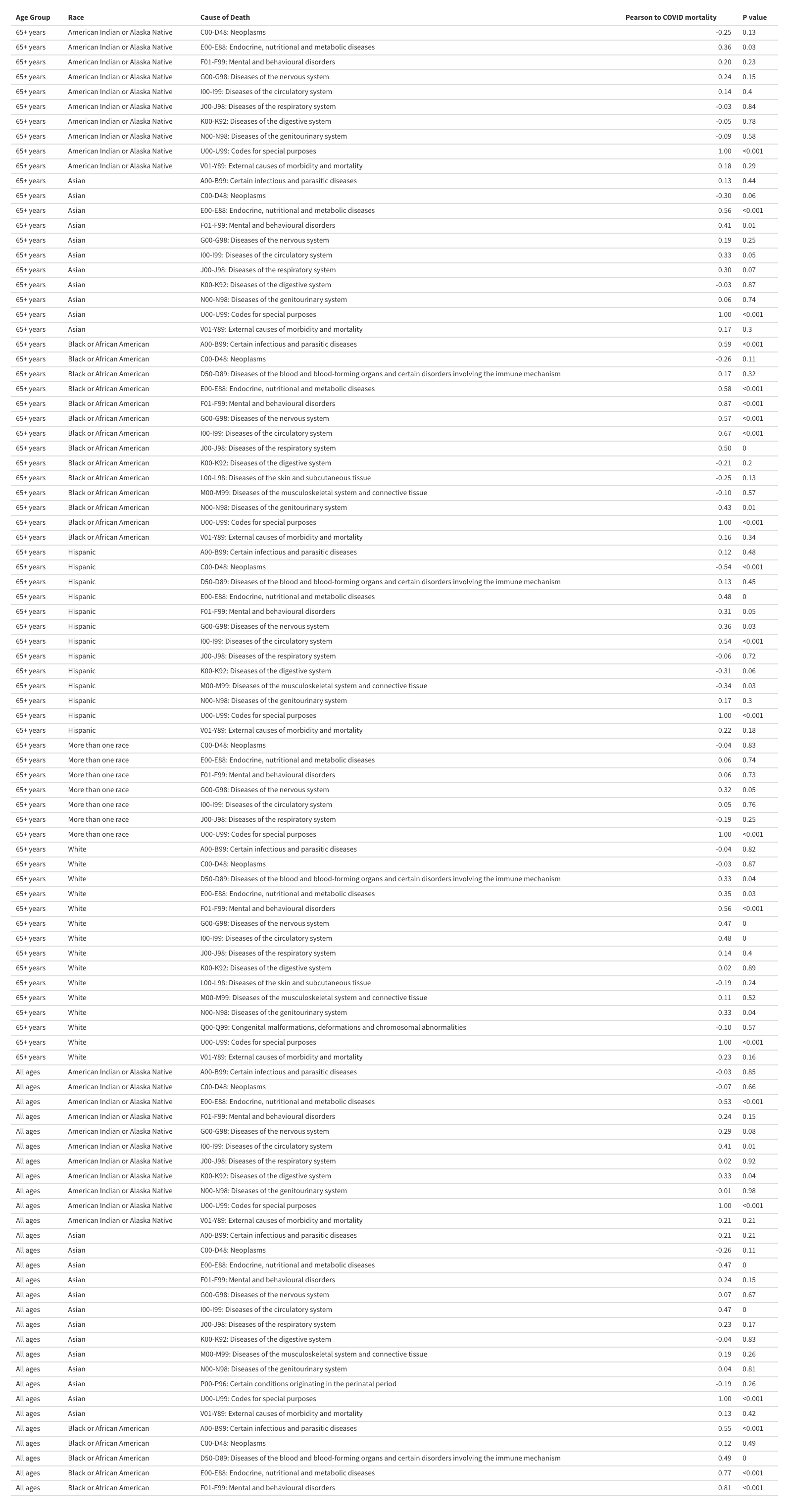

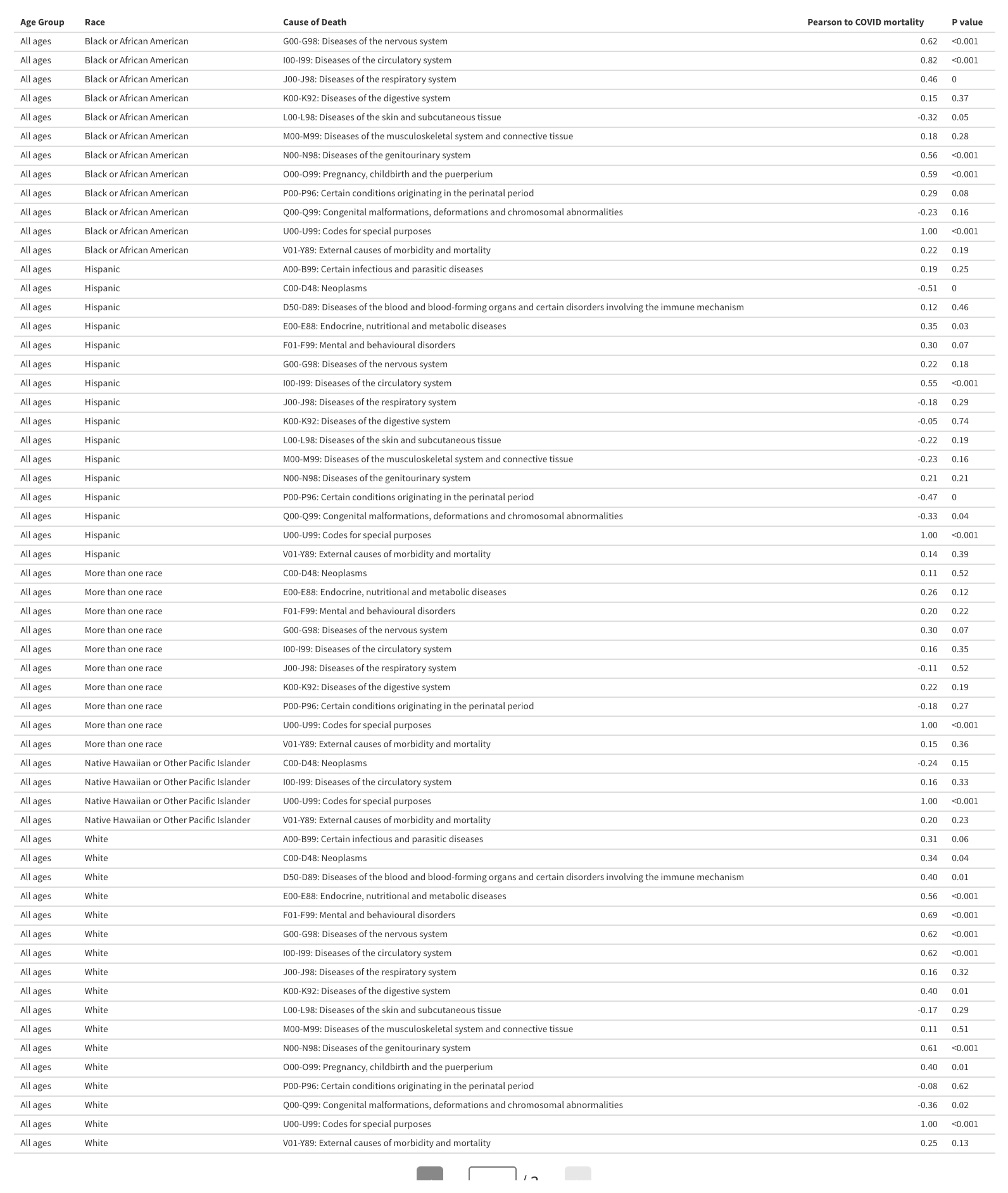

**Table S8.** Relative risks by age and race/ethnicity, pre-pandemic and pandemic periods.

|  | Pre-pandemic | Pandemic | Pre-pandemic | Pandemic | Pre-pandemic | Pandemic | Pre-pandemic | Pandemic |
| --- | --- | --- | --- | --- | --- | --- | --- | --- |
|  | <25 years | <25 years | 25-64 years | 25-64 years | 65+ years | 65+ years | All ages | All ages |
| American Indian or Alaska Native | 1.85 (1.80 - 1.90) | 2.16 (2.08 - 2.24) | 1.64 (1.62 - 1.66) | 2.10 (2.08 - 2.13) | 0.85 (0.84 - 0.86) | 0.85 (0.85 - 0.86) | 1.07 (1.06 - 1.08) | 1.22 (1.21 - 1.22) |
| Asian | 0.59 (0.58 - 0.61) | 0.57 (0.56 - 0.59) | 0.37 (0.37 - 0.38) | 0.38 (0.38 - 0.38) | 0.56 (0.56 - 0.56) | 0.57 (0.57 - 0.57) | 0.51 (0.51 - 0.51) | 0.52 (0.52 - 0.52) |
| Black or African American | 1.96 (1.94 - 1.97) | 2.31 (2.29 - 2.33) | 1.45 (1.45 - 1.46) | 1.58 (1.57 - 1.58) | 1.07 (1.07 - 1.07) | 1.11 (1.11 - 1.12) | 1.19 (1.19 - 1.19) | 1.26 (1.26 - 1.27) |
| Hispanic | 0.90 (0.89 - 0.90) | 1.04 (1.02 - 1.05) | 0.66 (0.66 - 0.67) | 0.80 (0.79 - 0.80) | 0.72 (0.71 - 0.72) | 0.79 (0.79 - 0.79) | 0.71 (0.71 - 0.71) | 0.80 (0.79 - 0.80) |
| More than one race | 0.67 (0.65 - 0.68) | 0.75 (0.73 - 0.77) | 0.52 (0.52 - 0.53) | 0.59 (0.58 - 0.60) | 0.41 (0.40 - 0.41) | 0.40 (0.40 - 0.41) | 0.44 (0.44 - 0.45) | 0.46 (0.46 - 0.46) |
| Native Hawaiian or Other Pacific Islander | 1.50 (1.41 - 1.60) | 1.79 (1.66 - 1.93) | 1.20 (1.17 - 1.23) | 1.50 (1.47 - 1.54) | 0.79 (0.77 - 0.80) | 0.78 (0.76 - 0.80) | 0.91 (0.90 - 0.92) | 1.00 (0.98 - 1.02) |
| White | 1.00 (0.99 - 1.01) | 1.00 (0.99 - 1.01) | 1.00 (1.00 - 1.00) | 1.00 (1.00 - 1.00) | 1.00 (1.00 - 1.00) | 1.00 (1.00 - 1.00) | 1.00 (1.00 - 1.00) | 1.00 (1.00 - 1.00) |

**Table S9.** Relative risks by age and race/ethnicity, pandemic period (pre-vaccine and post-vaccine periods).

|  | Pre-vaccine | Post-vaccine | Pre-vaccine | Post-vaccine | Pre-vaccine | Post-vaccine | Pre-vaccine | Post-vaccine |
| --- | --- | --- | --- | --- | --- | --- | --- | --- |
|  | <25 years | <25 years | 25-64 years | 25-64 years | 65+ years | 65+ years | All ages | All ages |
| American Indian or Alaska Native | 2.00 (1.89 - 2.13) | 2.25 (2.16 - 2.35) | 2.15 (2.11 - 2.18) | 2.08 (2.05 - 2.10) | 0.93 (0.92 - 0.95) | 0.82 (0.81 - 0.83) | 1.26 (1.25 - 1.27) | 1.19 (1.18 - 1.20) |
| Asian | 0.54 (0.52 - 0.57) | 0.59 (0.57 - 0.61) | 0.42 (0.41 - 0.42) | 0.36 (0.35 - 0.36) | 0.61 (0.61 - 0.62) | 0.55 (0.55 - 0.55) | 0.56 (0.56 - 0.56) | 0.50 (0.49 - 0.50) |
| Black or African American | 2.30 (2.26 - 2.34) | 2.32 (2.29 - 2.35) | 1.67 (1.66 - 1.67) | 1.52 (1.52 - 1.53) | 1.22 (1.22 - 1.23) | 1.07 (1.06 - 1.07) | 1.36 (1.35 - 1.36) | 1.21 (1.21 - 1.22) |
| Hispanic | 1.00 (0.98 - 1.02) | 1.06 (1.04 - 1.07) | 0.90 (0.89 - 0.90) | 0.74 (0.74 - 0.75) | 0.90 (0.90 - 0.91) | 0.74 (0.74 - 0.74) | 0.89 (0.89 - 0.90) | 0.74 (0.74 - 0.75) |
| More than one race | 0.72 (0.68 - 0.75) | 0.78 (0.75 - 0.80) | 0.58 (0.57 - 0.59) | 0.59 (0.58 - 0.60) | 0.40 (0.39 - 0.41) | 0.40 (0.40 - 0.41) | 0.46 (0.45 - 0.46) | 0.46 (0.46 - 0.47) |
| Native Hawaiian or Other Pacific Islander | 1.57 (1.37 - 1.79) | 1.93 (1.76 - 2.11) | 1.51 (1.46 - 1.58) | 1.49 (1.45 - 1.54) | 0.79 (0.76 - 0.83) | 0.78 (0.75 - 0.80) | 1.01 (0.98 - 1.03) | 1.00 (0.98 - 1.02) |
| White | 1.00 (0.98 - 1.02) | 1.00 (0.99 - 1.01) | 1.00 (1.00 - 1.00) | 1.00 (1.00 - 1.00) | 1.00 (1.00 - 1.00) | 1.00 (1.00 - 1.00) | 1.00 (1.00 - 1.00) | 1.00 (1.00 - 1.00) |

**Table S10.** Relative risks by age and race/ethnicity, by pandemic year, All ages.

|  | March 2015-February 2016 | March 2016-February 2017 | March 2017-February 2018 | March 2018-February 2019 | March 2019-February 2020 | March 2020-February 2021 | March 2021-February 2022 | March 2022-April 2023 |
| --- | --- | --- | --- | --- | --- | --- | --- | --- |
|  | All ages | All ages | All ages | All ages | All ages | All ages | All ages | All ages |
| American Indian or Alaska Native | 1.09 (1.08 - 1.11) | 1.08 (1.07 - 1.10) | 1.07 (1.05 - 1.08) | 1.07 (1.05 - 1.08) | 1.05 (1.04 - 1.07) | 1.27 (1.26 - 1.29) | 1.26 (1.25 - 1.28) | 1.13 (1.12 - 1.15) |
| Asian | 0.52 (0.52 - 0.53) | 0.51 (0.51 - 0.52) | 0.51 (0.51 - 0.52) | 0.51 (0.51 - 0.51) | 0.50 (0.50 - 0.51) | 0.56 (0.56 - 0.57) | 0.49 (0.49 - 0.50) | 0.51 (0.50 - 0.51) |
| Black or African American | 1.19 (1.18 - 1.19) | 1.19 (1.18 - 1.19) | 1.19 (1.19 - 1.19) | 1.19 (1.19 - 1.20) | 1.20 (1.19 - 1.20) | 1.36 (1.36 - 1.37) | 1.26 (1.26 - 1.27) | 1.19 (1.19 - 1.19) |
| Hispanic | 0.72 (0.71 - 0.72) | 0.71 (0.71 - 0.71) | 0.71 (0.70 - 0.71) | 0.70 (0.70 - 0.70) | 0.71 (0.70 - 0.71) | 0.91 (0.91 - 0.91) | 0.78 (0.78 - 0.79) | 0.72 (0.72 - 0.72) |
| More than one race | 0.44 (0.43 - 0.44) | 0.44 (0.43 - 0.45) | 0.46 (0.45 - 0.47) | 0.45 (0.44 - 0.46) | 0.44 (0.43 - 0.45) | 0.46 (0.45 - 0.46) | 0.46 (0.45 - 0.47) | 0.46 (0.46 - 0.47) |
| Native Hawaiian or Other Pacific Islander | 0.90 (0.87 - 0.94) | 0.91 (0.88 - 0.94) | 0.91 (0.88 - 0.94) | 0.91 (0.88 - 0.94) | 0.92 (0.89 - 0.95) | 0.99 (0.97 - 1.02) | 1.07 (1.04 - 1.10) | 0.95 (0.92 - 0.97) |
| White | 1.00 (1.00 - 1.00) | 1.00 (1.00 - 1.00) | 1.00 (1.00 - 1.00) | 1.00 (1.00 - 1.00) | 1.00 (1.00 - 1.00) | 1.00 (1.00 - 1.00) | 1.00 (1.00 - 1.00) | 1.00 (1.00 - 1.00) |

### **Table S11.** Relative risks by age and race/ethnicity, by pandemic year, Ages <25 years.

|  | March 2015-February 2016 | March 2016-February 2017 | March 2017-February 2018 | March 2018-February 2019 | March 2019-February 2020 | March 2020-February 2021 | March 2021-February 2022 | March 2022-April 2023 |
| --- | --- | --- | --- | --- | --- | --- | --- | --- |
|  | <25 years | <25 years | <25 years | <25 years | <25 years | <25 years | <25 years | <25 years |
| American Indian or Alaska Native | 1.83 (1.71 - 1.95) | 1.83 (1.72 - 1.95) | 1.86 (1.74 - 1.99) | 1.82 (1.70 - 1.95) | 1.95 (1.82 - 2.08) | 2.03 (1.91 - 2.17) | 2.16 (2.03 - 2.30) | 2.27 (2.14 - 2.40) |
| Asian | 0.60 (0.57 - 0.63) | 0.59 (0.56 - 0.62) | 0.59 (0.57 - 0.62) | 0.60 (0.57 - 0.63) | 0.58 (0.56 - 0.61) | 0.53 (0.51 - 0.56) | 0.58 (0.56 - 0.61) | 0.60 (0.57 - 0.62) |
| Black or African American | 1.90 (1.87 - 1.94) | 1.92 (1.88 - 1.96) | 1.91 (1.87 - 1.94) | 1.97 (1.93 - 2.01) | 2.09 (2.05 - 2.13) | 2.30 (2.25 - 2.34) | 2.33 (2.29 - 2.38) | 2.30 (2.26 - 2.34) |
| Hispanic | 0.88 (0.86 - 0.90) | 0.87 (0.85 - 0.89) | 0.88 (0.87 - 0.90) | 0.89 (0.87 - 0.91) | 0.97 (0.95 - 0.99) | 1.00 (0.98 - 1.03) | 1.03 (1.01 - 1.05) | 1.07 (1.05 - 1.09) |
| More than one race | 0.60 (0.57 - 0.63) | 0.60 (0.57 - 0.64) | 0.70 (0.67 - 0.74) | 0.69 (0.66 - 0.73) | 0.75 (0.71 - 0.78) | 0.72 (0.68 - 0.75) | 0.75 (0.71 - 0.78) | 0.79 (0.76 - 0.82) |
| Native Hawaiian or Other Pacific Islander | 1.23 (1.05 - 1.45) | 1.51 (1.31 - 1.74) | 1.45 (1.25 - 1.68) | 1.63 (1.41 - 1.88) | 1.79 (1.56 - 2.06) | 1.51 (1.30 - 1.75) | 1.89 (1.66 - 2.15) | 1.97 (1.75 - 2.22) |
| White | 1.00 (0.98 - 1.02) | 1.00 (0.98 - 1.02) | 1.00 (0.98 - 1.02) | 1.00 (0.98 - 1.02) | 1.00 (0.98 - 1.02) | 1.00 (0.98 - 1.02) | 1.00 (0.98 - 1.02) | 1.00 (0.98 - 1.02) |

### **Table S12.** Relative risks by age and race/ethnicity, by pandemic year, Ages 25-64 years.

|  | March 2015-February 2016 | March 2016-February 2017 | March 2017-February 2018 | March 2018-February 2019 | March 2019-February 2020 | March 2020-February 2021 | March 2021-February 2022 | March 2022-April 2023 |
| --- | --- | --- | --- | --- | --- | --- | --- | --- |
|  | 25-64 years | 25-64 years | 25-64 years | 25-64 years | 25-64 years | 25-64 years | 25-64 years | 25-64 years |
| American Indian or Alaska Native | 1.63 (1.59 - 1.67) | 1.60 (1.57 - 1.64) | 1.62 (1.59 - 1.66) | 1.68 (1.64 - 1.72) | 1.68 (1.64 - 1.71) | 2.18 (2.14 - 2.22) | 2.13 (2.10 - 2.17) | 2.01 (1.98 - 2.05) |
| Asian | 0.38 (0.37 - 0.39) | 0.37 (0.37 - 0.38) | 0.37 (0.36 - 0.38) | 0.37 (0.37 - 0.38) | 0.37 (0.36 - 0.38) | 0.42 (0.41 - 0.43) | 0.36 (0.35 - 0.36) | 0.37 (0.36 - 0.37) |
| Black or African American | 1.44 (1.43 - 1.45) | 1.44 (1.43 - 1.45) | 1.45 (1.44 - 1.46) | 1.46 (1.45 - 1.47) | 1.47 (1.46 - 1.48) | 1.67 (1.67 - 1.68) | 1.55 (1.55 - 1.56) | 1.51 (1.50 - 1.52) |
| Hispanic | 0.67 (0.66 - 0.67) | 0.66 (0.65 - 0.66) | 0.66 (0.66 - 0.67) | 0.66 (0.66 - 0.67) | 0.67 (0.67 - 0.68) | 0.92 (0.91 - 0.92) | 0.79 (0.78 - 0.79) | 0.72 (0.71 - 0.72) |
| More than one race | 0.50 (0.48 - 0.52) | 0.50 (0.48 - 0.51) | 0.55 (0.53 - 0.56) | 0.54 (0.52 - 0.55) | 0.54 (0.53 - 0.56) | 0.58 (0.57 - 0.60) | 0.58 (0.56 - 0.59) | 0.61 (0.59 - 0.62) |
| Native Hawaiian or Other Pacific Islander | 1.19 (1.13 - 1.26) | 1.13 (1.07 - 1.19) | 1.20 (1.14 - 1.26) | 1.22 (1.16 - 1.29) | 1.24 (1.18 - 1.31) | 1.52 (1.45 - 1.58) | 1.61 (1.55 - 1.67) | 1.40 (1.35 - 1.46) |
| White | 1.00 (1.00 - 1.00) | 1.00 (1.00 - 1.00) | 1.00 (1.00 - 1.00) | 1.00 (1.00 - 1.00) | 1.00 (1.00 - 1.00) | 1.00 (1.00 - 1.00) | 1.00 (1.00 - 1.00) | 1.00 (1.00 - 1.00) |

### **Table S13.** Relative risks by age and race/ethnicity, by pandemic year, Ages ≥65 years.

|  | March 2015-February 2016 | March 2016-February 2017 | March 2017-February 2018 | March 2018-February 2019 | March 2019-February 2020 | March 2020-February 2021 | March 2021-February 2022 | March 2022-April 2023 |
| --- | --- | --- | --- | --- | --- | --- | --- | --- |
|  | 65+ years | 65+ years | 65+ years | 65+ years | 65+ years | 65+ years | 65+ years | 65+ years |
| American Indian or Alaska Native | 0.88 (0.86 - 0.90) | 0.88 (0.86 - 0.90) | 0.84 (0.83 - 0.86) | 0.83 (0.81 - 0.84) | 0.80 (0.79 - 0.82) | 0.93 (0.92 - 0.95) | 0.88 (0.86 - 0.89) | 0.77 (0.76 - 0.79) |
| Asian | 0.57 (0.56 - 0.57) | 0.56 (0.55 - 0.56) | 0.56 (0.55 - 0.56) | 0.55 (0.55 - 0.56) | 0.55 (0.54 - 0.55) | 0.61 (0.61 - 0.62) | 0.55 (0.54 - 0.55) | 0.55 (0.55 - 0.56) |
| Black or African American | 1.07 (1.07 - 1.08) | 1.07 (1.07 - 1.08) | 1.07 (1.07 - 1.08) | 1.07 (1.07 - 1.08) | 1.07 (1.06 - 1.07) | 1.22 (1.22 - 1.23) | 1.11 (1.10 - 1.11) | 1.03 (1.03 - 1.04) |
| Hispanic | 0.73 (0.72 - 0.73) | 0.72 (0.72 - 0.72) | 0.72 (0.71 - 0.72) | 0.71 (0.70 - 0.71) | 0.71 (0.71 - 0.71) | 0.90 (0.90 - 0.91) | 0.77 (0.77 - 0.78) | 0.71 (0.71 - 0.71) |
| More than one race | 0.41 (0.40 - 0.42) | 0.41 (0.40 - 0.42) | 0.42 (0.41 - 0.43) | 0.41 (0.40 - 0.42) | 0.40 (0.39 - 0.41) | 0.40 (0.39 - 0.41) | 0.40 (0.39 - 0.41) | 0.40 (0.39 - 0.41) |
| Native Hawaiian or Other Pacific Islander | 0.79 (0.75 - 0.83) | 0.80 (0.77 - 0.85) | 0.79 (0.75 - 0.82) | 0.77 (0.73 - 0.81) | 0.77 (0.74 - 0.81) | 0.79 (0.76 - 0.83) | 0.82 (0.78 - 0.85) | 0.74 (0.71 - 0.77) |
| White | 1.00 (1.00 - 1.00) | 1.00 (1.00 - 1.00) | 1.00 (1.00 - 1.00) | 1.00 (1.00 - 1.00) | 1.00 (1.00 - 1.00) | 1.00 (1.00 - 1.00) | 1.00 (1.00 - 1.00) | 1.00 (1.00 - 1.00) |
